# Ensemble SHAP Aggregation and Attribution Variability in Clinical Machine Learning: A COVID-19 Mortality Study

**DOI:** 10.64898/2026.09.07.26362459

**Authors:** Na Dai, Ruben K. Briceno, Alex N. Castaneda, Miguel A. Tresierra, Maribel L. Esteban, Moises E. Rosas, Rene C. Hinojosa

## Abstract

**Objective:** To combine performance-weighted ensemble SHAP aggregation with resampling-based assessment of feature-importance variability and patient-level attribution alignment for interpreting COVID-19 mortality predictions.

**Methods:** We analyzed a prospective cohort of 1,857 patients hospitalized with COVID-19 at two hospitals in Peru. Ten predictive algorithms were evaluated using five-fold cross-validation, and their mean AUROC values determined their attribution-aggregation weights. Within each model and training trial, signed SHAP values were normalized by the mean total absolute attribution across evaluation patients before performance-weighted aggregation. Feature importance was summarized across 30 resampled training trials using means and normal-approximation 95% confidence intervals. Patient-level attribution alignment was assessed using cosine similarity between signed feature-attribution vectors for corresponding patients, with whole-trial patient-correspondence randomization and Benjamini–Hochberg correction. Subgroup observations were reweighted toward the evaluated population’s feature distributions for secondary comparisons of existing absolute SHAP values.

**Results:** Among the 1,857 included patients, 982 (52.9%) died during hospitalization. Random forest achieved the highest mean AUROC (0.925 *±* 0.010), followed by AdaBoost (0.919 *±* 0.017) and logistic regression (0.918 *±* 0.007), with variability reported as the SEM. Attribution profiles were more similar across repeated training trials of the same algorithm (mean Pearson *r* = 0.783; SEM, 0.004) than across algorithms within the same trial (mean Pearson *r* = 0.369; SEM, 0.006). The largest normalized attribution magnitudes were observed for dexamethasone use at home without oxygen support (6.186%; 95% CI, 5.795–6.577), PaO_2_/FiO_2_ ratio (3.014%; 95% CI, 2.787–3.242), shortness of breath (2.774%; 95% CI, 2.592–2.956), and FiO_2_ (2.560%; 95% CI, 2.284–2.836). Features differed in patient-level attribution alignment, indicating that importance magnitude and robustness provided complementary information. Among 1,763 eligible subgroup–feature comparisons, 64 had nominal one-sided *p <* 0.05, although none remained below 0.05 after Benjamini–Hochberg adjustment.

**Conclusions:** Algorithms with similar predictive performance produced substantially different feature-attribution profiles. Normalized performance-weighted SHAP aggregation provided a representative relative-importance summary across algorithms, while resampling-based alignment assessment qualified the consistency of individual feature contributions. The resulting attribution patterns describe fitted-model behavior and should not be interpreted as causal clinical effects.

## 1 Introduction

Clinical prediction models can estimate patient risk, but interpreting that risk requires understanding which clinical features contribute to it. SHapley Additive exPlanations (SHAP) support this interpretation through feature attribution [14]. However, well-performing models may assign different importance to the same variables [4], potentially leading to different clinical interpretations. This motivates assessing the representativeness and robustness of feature-importance profiles across modeling algorithms and training samples.

To reduce dependence on individual models, we draw on the rationale of ensemble learning: combining information from multiple fitted models to construct a representative summary. Previous work has demonstrated that aggregating feature-based explanations can reduce their sensitivity [1]. Building on this rationale, we use ensemble SHAP aggregation to integrate attributions from diverse predictive algorithms using performance-based weights, providing a representative feature-importance profile for clinical interpretation.

However, an aggregated profile can conceal differences in feature-level robustness. Features with similar average importance may contribute consistently across fits or receive large contributions in only a subset of models and trials. We therefore complement aggregation with resampling-based robustness analysis to characterize the variability underlying each importance estimate. Considering magnitude and robustness together distinguishes prominent but variable contributions from those supported more consistently.

This assessment can help place model findings within a coherent clinical account. Alignment with established knowledge supports clinical plausibility, while less familiar patterns may suggest hypotheses about mortality-associated physiology. Robustness helps qualify the support for these interpretations, which describe model behavior and do not independently establish biological causality [11].

We propose an integrated approach that combines performance-weighted ensemble SHAP aggregation with resampling-based robustness assessment to support clinical interpretation of feature importance. We investigate this approach in a prospective cohort of 1,857 patients hospitalized with COVID-19 at two hospitals in Peru. Building on previous applications of machine learning to mortality prediction and clinical feature identification [23], we aggregate SHAP attributions from ten predictive algorithms, assess variability across repeated training trials, and examine subgroup patterns through distribution-adjusted reweighting.

This study makes three contributions:

- An integrated workflow that pairs a representative ensemble feature-importance profile with assessment of the robustness of its underlying contributions.
- An empirical evaluation of how feature importance varies across predictive algorithms and repeated training trials.
- A clinical analysis of COVID-19 mortality that interprets feature importance alongside robustness, including distribution-adjusted subgroup comparisons.

## 2 Related Work

### 2.1 Attribution Variability Across Models and Data

#### Variation across models

Different predictive models can assign different importance to the same clinical features, even when evaluated on the same patients. Fisher et al. introduced model class reliance to characterize the range of variable importance across well-performing models [4]. Extending this perspective to Shapley-based importance, Ning et al. developed ShapleyVIC to pool importance estimates across models and quantify their uncertainty, including in a clinical mortality application [16]. Laberge et al. identified local and global importance relationships supported across a Rashomon set of similarly performing models, allowing features to remain unordered when models disagree [12]. Collectively, these approaches characterize importance ranges, pooled estimates, and attribution consensus across plausible models. Our study builds on this foundation by comparing SHAP attributions across heterogeneous clinical prediction algorithms and integrating them into a performance-weighted profile.

#### Variation among training sets

Feature importance can also change when the same modeling approach is trained on different samples, even when explanations are evaluated on the same patients. Yasodhara et al. demonstrated sensitivity of global SHAP importance to input noise, random seeds, and hyperparameter changes [24]. Addressing sampling variability more directly, Xiang et al. [22] used bootstrap attribution distributions to construct RoSHAP, a ranking criterion incorporating attribution frequency, strength, and stability. These studies characterize perturbation sensitivity and distributional variation, but global importance and ranking summaries can obscure changes in signed contributions across individual observations. We examine these changes by comparing feature-specific attribution patterns across the same patients after repeated resampling and retraining, complementing assessments based on average importance and ranking consistency.

### 2.2 Explanation Aggregation and Uncertainty Assessment

Aggregation offers a means of summarizing explanations beyond individual fitted models. Bhatt et al. developed explanation-aggregation procedures to reduce explanation sensitivity and improve other explanation properties [1]. More directly related to model combination, Haghish’s WMSHAP software provides weighted SHAP summaries and confidence intervals across fitted models [7]. These approaches establish a basis for combining attributions, but an aggregated estimate alone does not reveal whether its underlying patient-level contributions remain consistent across training samples. Our study therefore pairs performance-weighted SHAP aggregation with resampling-based assessment of feature-level robustness.

Previous work has also proposed Shapley-based methods to assess distinct sources of uncertainty. Li et al. developed DistDeepSHAP to quantify reference-dependent attribution variation by sampling background observations for a fixed model and input [13]. Johnsen et al. introduced Sub-SAGE to measure global feature importance through reductions in expected prediction loss, using paired bootstrap resampling of independent test observations to estimate sampling uncertainty conditional on a fitted model [9]. In these uncertainty assessments, the predictive model is held fixed: the former examines variation across reference observations, whereas the latter estimates sampling uncertainty from evaluation data. Our study complements these assessments by examining SHAP attribution variability across predictive algorithms and resampled training datasets.

### 2.3 Clinical Applications and Subgroup Interpretation

Feature-importance analysis has been used to connect COVID-19 mortality predictions with clinically interpretable patient characteristics. Among the early studies in this area, Yadaw et al. [23] identified age, minimum oxygen saturation, and encounter type as informative predictors of mortality. Moving from predictor identification to attribution analysis, Booth et al. [2] used SHAP to examine how laboratory measurements contributed to mortality predictions from a support-vector-machine model. Smith and Alvarez [18] further examined global and patient-level Shapley explanations across machine-learning models, relating predicted mortality to individual patient characteristics. These studies demonstrate how feature interpretation can characterize mortality-associated patterns, motivating further assessment of how those interpretations depend on model choice and training data.

Beyond COVID-19, feature interpretation has also been applied in other clinical settings. Lundberg et al. [15] developed a system that paired intraoperative hypoxaemia predictions with explanations of contributing factors, examined whether those explanations agreed with clinical knowledge, and evaluated anaesthesiologists’ predictions with system assistance. Incorporating variability assessment into clinical modeling, Huang and Huang [8] examined distributions of predictive performance and feature-gain statistics across repeated training and test splits, alongside SHAP-based interpretation of heart-disease predictors. Their resampling summaries concerned performance and gain statistics rather than patient-level SHAP consistency. Together, these studies demonstrate the clinical value of interpreting feature contributions while considering the variability underlying model-derived findings.

Clinical research has also begun to examine whether model explanations remain stable across modeling conditions. Riley and Collins [17] used bootstrap model redevelopment to evaluate instability in clinical predictions and cautioned that post hoc explanations may be misleading when the underlying models are unstable. Connecting resampling with subgroup assessment, Ellis and Polberg-Riener [3] evaluated how female-targeted resampling affected predictive utility, calibration, and explanation stability across clinical datasets and model families, measuring stability through feature-importance drift and partial-dependence divergence. Focusing directly on SHAP explanations, Guillén and Frias-Martinez [5] assessed their coherence and stability across model architectures, disease-stage boundaries, and diagnostic and prognostic tasks in Alzheimer’s disease.

A related line of research has compared attribution profiles across patient groups. Wang et al. [21] examined SHAP importance across sex, race and ethnicity, and insurance subgroups in an emergency-department prediction model, identifying shared and subgroup-specific predictors. Such comparisons use the observed feature distributions within each subgroup, so apparent differences in importance may partly reflect differences in subgroup composition.

Taken together, existing studies have examined prediction stability, explanation consistency across modeling settings, and SHAP differences across observed patient groups. However, they do not jointly connect an aggregated attribution profile with the consistency of each feature’s signed contributions for the same patients across resampled training sets or assess subgroup differences after accounting for differences in feature distributions. Our study brings these elements together through ensemble SHAP aggregation, resampling-based robustness assessment, and distribution-adjusted subgroup comparisons.

## 3 Methods

### 3.1 Study Design and Population

We conducted a prospective cohort study of patients hospitalized with coronavirus disease 2019 (COVID-19) in the inpatient wards of Hospital de Alta Complejidad Virgen de la Puerta and Hospital Víctor Lazarte Echegaray (EsSalud) in Peru between March 1 and December 31, 2020. Patients were eligible if they were admitted for inpatient hospitalization with COVID-19 at either participating hospital during the study period. Of 2,000 admitted patients, 143 with documented psychiatric conditions were excluded, leaving 1,857 patients in the analysis.

Clinical data were collected prospectively from enrollment and throughout hospitalization until recovery and discharge or in-hospital death. COVID-19 was diagnosed using molecular testing, antibody or antigen assays, or the treating clinician’s assessment based on the clinical presentation. The primary outcome was in-hospital mortality.

### 3.2 Feature Construction and Preprocessing

A total of 279 candidate clinical variables were extracted. These covered demographics, history of presenting illness, ABO blood type, past medical history, evidence of bacterial infection, symptoms, laboratory and physiologic measurements, imaging findings, treatments, and medications. Continuous measurements were retained numerically and, where clinically relevant, were also represented using predefined categories, such as *PaO*_2_/*FiO*_2_ *>* 300 and *PaO*_2_/*FiO*_2_ between 200 and 300. Feature encoding produced 283 model inputs.

For the reported analyses, missing values were imputed by *k*-nearest-neighbor imputation using two neighbors with uniform weights. Predictor values were subsequently transformed to the range [0, 1]. Imputation and scaling were fitted on each trial-specific training subset and then applied without refitting to its evaluation observations. Vaccination indicators were retained during model fitting but omitted from the reported feature-importance results because only one patient had a recorded COVID-19 vaccination, providing insufficient exposure variation for meaningful interpretation.

### 3.3 Predictive Models and Cross-Validation

Ten predictive algorithms were evaluated: logistic regression (LR), linear support vector machine (Linear SVM), support vector machine with a radial basis function kernel (RBF SVM), random forest, decision tree, multilayer perceptron (MLP), AdaBoost, Gaussian naïve Bayes (NB), quadratic discriminant analysis (QDA), and a convolutional neural network (CNN). Logistic regression used the SAGA solver with an elastic-net penalty, an *ℓ*_1_ ratio of 0.5, and a tolerance of 0.02. Both SVM models used *C* = 0.5. The random forest contained 300 trees, and the MLP used *α* = 3 with a maximum of 100 iterations. Other scikit-learn models used default hyperparameters unless otherwise specified. The CNN contained two one-dimensional convolutional layers with 256 and 64 filters, respectively, each followed by average pooling; the resulting representation was flattened and passed through a 32-unit rectified-linear dense layer and a one-unit sigmoid output layer.

Predictive performance was evaluated by five-fold cross-validation on the full cohort. The area under the receiver operating characteristic curve (AUROC), accuracy, sensitivity, specificity, positive predictive value (PPV), and negative predictive value (NPV) were summarized as the mean and standard error of the mean (SEM) across folds. Let *A_k_* denote the mean cross-validated AUROC for algorithm *k*. Its attribution weight was

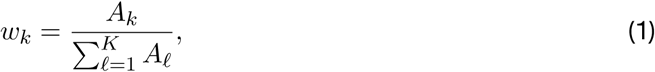

where *K* = 10 and 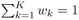. These weights were fixed for all subsequent attribution trials. Because the cross-validation used the full cohort, observations later included in the attribution evaluation subsets also contributed to weight estimation.

### 3.4 Resampled Training Trials

The attribution analysis comprised 30 training trials per algorithm, organized into three evaluation batches of ten trials. Each batch used a fixed evaluation subset containing 10% of the cohort (186 patients). For each trial, 30% of the available observations were randomly selected as the training subset, without replacement within that trial and using a trial-specific random seed. A patient could therefore appear in multiple trials but no more than once in a given training subset. The training and evaluation subsets were selected independently; consequently, evaluation observations were not guaranteed to be excluded from a trial’s training subset.

Within an evaluation batch, all ten algorithms and all ten training trials were explained on the same patients in the same order. This correspondence enabled patient-level comparison of attributions across fitted models. The three batches contained different evaluation subsets, so patient-level comparisons were restricted to trials within the same batch.

### 3.5 SHAP Attribution

SHapley Additive exPlanations (SHAP) were used to describe the contribution of each feature to an individual model output. For a fitted model *f* and patient *x*, the SHAP representation is

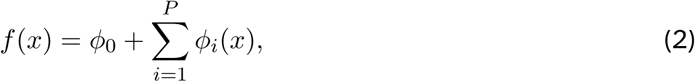

where *ϕ*_0_ is the explainer baseline, *ϕ_i_*(*x*) is the signed attribution for feature *i*, and *P* = 283 is the number of encoded inputs. Positive and negative values indicate contributions that increase or decrease the explained output relative to its baseline.

TreeExplainer was used for the decision-tree and random-forest models and was initialized without an explicit background dataset. KernelExplainer was used for the other algorithms. For KernelExplainer, the corresponding evaluation matrix served both as the background reference and as the observations to be explained. The non-tree scikit-learn models were explained using their `predict_proba` output, while the CNN was explained using its `predict` output. The retained class index was 1 for the non-CNN models and 0 for the single-output CNN; both represented the mortality output used in the analysis. KernelExplainer used the identity link. No additional transformation was applied to the raw SHAP values before the normalization described below.

### 3.6 Normalized Ensemble SHAP Aggregation

Raw SHAP magnitudes can differ across algorithms because their fitted functions, output representations, and explainer baselines differ. We therefore standardized the overall attribution magnitude of each model–trial before combining models. For algorithm *k*, batch *b*, and trial *t*, the attribution budget was defined as

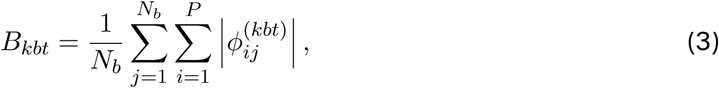

where *N_b_* = 186 is the number of evaluation patients in batch *b*. The normalized signed attribution was

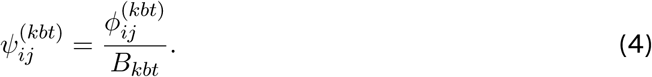

This operation set the mean patient-level total absolute attribution to one within each model–trial while retaining the relative magnitudes and signs of its patient-level contributions.

The normalized attributions were then aggregated across algorithms using the fixed AUROC weights:

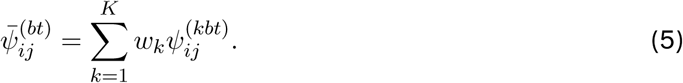

This quantity is an empirical performance-weighted summary of attributions from heterogeneous fitted models. It is not the exact SHAP decomposition of a single ensemble prediction function.

For feature *i* in trial *t* of batch *b*, ensemble importance was defined as

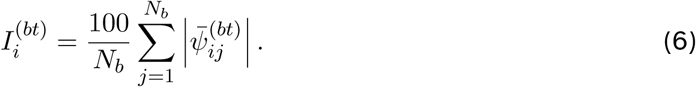

Multiplication by 100 expresses the result relative to the standardized attribution budget. Because signed contributions were combined across models before their absolute values were taken, feature importances need not sum to 100. For each feature, the mean, standard deviation, and SEM were calculated from the 30 trial-specific estimates. Normal-approximation 95% confidence intervals were calculated as the mean *±*1.96*×* SEM, with negative lower limits truncated at zero because importance is nonnegative.

### 3.7 Attribution Variation Across Algorithms and Training Trials

To compare global attribution profiles, model-specific normalized importance was calculated as

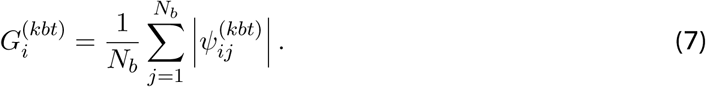

Pearson correlations across all encoded features were then calculated for two types of compaison: the same algorithm across unique pairs of training trials within an evaluation batch, and different algorithms within the same trial and batch. Each comparison comprised 1,350 profile pairs: 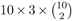 for the former and 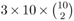 for the latter. Correlations were summarized by their mean and SEM.

### 3.8 Patient-Level Attribution Alignment

Feature importance describes average attribution magnitude but does not indicate whether a feature contributes similarly for the same patients across training trials. For each feature *i*, batch *b*, and trial *t*, we therefore formed the signed ensemble-attribution vector

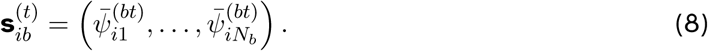

For each unique pair of trials *a < b* within the same batch, alignment was quantified by cosine similarity:

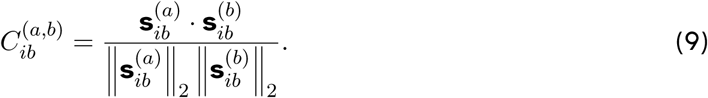

Cosine similarity ranges from *−*1 to 1. Positive values indicate that the signed attribution patterns tend to align across corresponding patients, values near zero indicate little patient-specific alignment, and negative values indicate opposing patterns. The reported alignment for each feature was the mean across the 45 unique trial pairs in each batch and then across the three batches, giving 135 pairwise comparisons per feature.

A one-sided patient-correspondence randomization test evaluated whether the observed mean cosine alignment exceeded that expected without consistent patient matching across trials. For each of 2,000 randomizations, the patient labels of each whole trial vector were independently permuted within its evaluation batch. This preserved the attribution values and within-trial vector structure while breaking correspondence for the same patients across trials. The complete alignment statistic was recalculated after each randomization. If 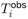 is the observed mean alignment and 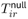 is its value in randomization *r*, the p-value was

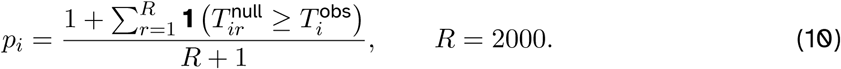

The resulting p-values were adjusted across all 283 encoded features using the Benjamini–Hochberg false-discovery-rate procedure. Table 3 reports the adjusted *q*-values. These tests concern patient-specific alignment of signed model attributions; they do not test association with mortality, importance magnitude, or a clinical effect.

### 3.9 Visualization of Signed Ensemble Attributions

For the supplementary beeswarm plots, normalized ensemble attributions were averaged across the ten trials within each fixed evaluation batch and then pooled across the three batches, yielding 558 plotted evaluation observations per feature. The horizontal axis represents signed normalized ensemble attribution, expressed on the same percentage scale as Equation 6. Feature color represents the within-batch percentile rank of the observed feature value, allowing visual comparison despite preprocessing differences. Separate plots show up to the 20 most important features within each of 16 clinical categories; categories containing fewer than 20 reported features show all available features.

### 3.10 Distribution-Adjusted Subgroup Analysis

Distribution-adjusted subgroup analysis used the normalized ensemble attributions generated for Table 4. The archived attribution data comprised three evaluation batches of 186 observations each, yielding 558 pooled evaluation observations. Original cohort identifiers were not retained across batches; the pooled records are therefore described as evaluation observations rather than unique patients. For every subgroup *G*, the pooled evaluation reference *R* comprised all 558 available evaluation observations, including observations belonging to *G*.

Evaluation observations were grouped by age 50 years or older, sex, ABO blood group, asthma or chronic obstructive pulmonary disease, metabolic or cardiovascular disease, kidney disease, chronic viral or liver disease, and the presence of at least two documented comorbidities. Metabolic or cardiovascular disease comprised type 2 diabetes, hypertension, ischemic heart disease, or obesity. Kidney disease comprised acute kidney injury or chronic kidney disease stages 1–5. Chronic viral or liver disease comprised HIV, hepatitis C, compensated or decompensated cirrhosis, or hepatocellular carcinoma.

For feature *i* and evaluation observation *j* in batch *b*, patient-level attribution magnitude was calculated by averaging the absolute normalized ensemble attribution across the ten training trials in that batch:

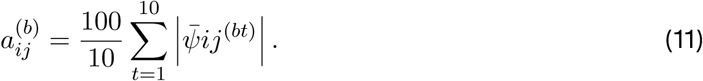

The batch superscript is omitted below when observations from the three batches are pooled. Pooled-reference importance was

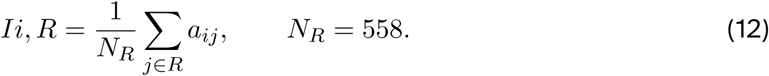

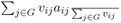 where *v_ij_* is the feature-specific weight. Absolute and relative differences were calculated as

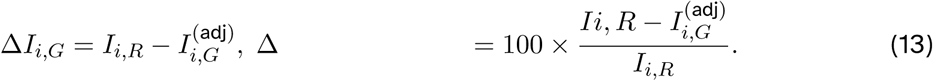

Positive differences therefore indicated greater attribution magnitude in the pooled evaluation reference, whereas negative differences indicated greater magnitude in the distribution-adjusted subgroup.

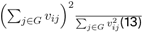 was below 10. Features with zero patient-level attribution variance, for which the Student’s *t* statistic was undefined, were also excluded from inference.

For eligible features, nominal *p*-values were calculated from the patient-level absolute SHAP magnitudes *a_ij_* using a weighted, pooled-variance Student’s *t* test. Let 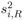 denote the variance of *a_ij_* in the pooled evaluation reference and let 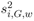 denote the feature-weighted variance in subgroup *G*. The pooled variance and test statistic were

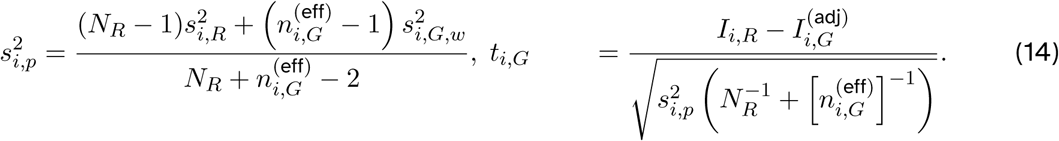

The degrees of freedom were 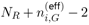, and the reported one-sided tail probability in the direction of the observed difference was

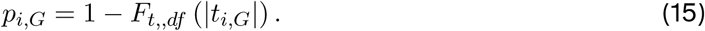

The test was conducted on attribution magnitudes rather than signed SHAP values because Table 4 compares feature importance. Because subgroup observations also occurred in the pooled evaluation reference and uncertainty in the estimated reweighting weights was not propagated, these *p*-values were treated as nominal and exploratory.

All 283 encoded attributes entered the subgroup calculations. Attributes defining a particular subgroup and attributes failing the balance or effective-sample-size criteria were excluded from inference only for the affected subgroup. Subgroups containing fewer than 20 evaluation observations were summarized descriptively without inferential values. Within each subgroup, Table 4 displays up to 20 attributes with the largest absolute differences; selection was based on *|*Δ*I|*, rather than on the nominal *p*-value. Benjamini–Hochberg adjustments were calculated both within each subgroup and across all eligible subgroup–attribute comparisons and were retained in the complete supplementary data as sensitivity summaries. These comparisons describe differences in model attribution after marginal distribution adjustment and do not establish subgroup-specific biological or treatment effects.

## 4 Results

### 4.1 Performance on COVID-19 Mortality Prediction

Among the 1,857 included patients, 982 (52.9%) died during hospitalization and 875 (47.1%) survived to discharge. Table 1 summarizes predictive performance under fivefold cross-validation. Seven algorithms achieved mean AUROC values between 0.907 and 0.925. Random forest had the highest mean AUROC (0.925 *±* 0.010 SEM), followed by AdaBoost (0.919 *±* 0.017), logistic regression (0.918 *±* 0.007), the multilayer perceptron (0.915 *±* 0.012), linear SVM (0.913 *±* 0.013), the CNN (0.911 *±* 0.006), and RBF SVM (0.907 *±* 0.008). Gaussian naïve Bayes, decision tree, and quadratic discriminant analysis achieved lower mean AUROCs of 0.854, 0.775, and 0.768, respectively.

**Table 1:** COVID-19 mortality prediction performance (mean *±* SEM^1^) across ten predictive algorithms under fivefold cross-validation.

| Algorithm | AUROC<sup>5</sup> | Accuracy | Sensitivity | Specificity | PPV<sup>6</sup> | NPV<sup>7</sup> |
| --- | --- | --- | --- | --- | --- | --- |
| LR<sup>3</sup> | 0.918±0.007 | 0.841±0.009 | 0.853±0.025 | 0.827±0.032 | 0.848±0.020 | 0.835±0.020 |
| Linear SVM ($C = 0.5$) | 0.913±0.013 | 0.836±0.013 | 0.848±0.021 | 0.822±0.032 | 0.843±0.023 | 0.829±0.017 |
| RBF SVM ($C = 0.5$) | 0.907±0.008 | 0.815±0.008 | 0.804±0.023 | 0.829±0.027 | 0.841±0.019 | 0.790±0.016 |
| Random forest (300 trees)<sup>4</sup> | 0.925±0.010 | 0.844±0.010 | 0.828±0.021 | 0.863±0.023 | 0.872±0.017 | 0.818±0.016 |
| Decision tree<sup>4</sup> | 0.775±0.017 | 0.776±0.017 | 0.799±0.021 | 0.750±0.042 | 0.783±0.028 | 0.769±0.016 |
| MLP ($\alpha = 3$, maximum 100 iterations)<sup>4</sup> | 0.915±0.012 | 0.827±0.012 | 0.806±0.052 | 0.850±0.043 | 0.860±0.029 | 0.799±0.035 |
| AdaBoost<sup>4</sup> | 0.919±0.017 | 0.850±0.017 | 0.861±0.010 | 0.838±0.030 | 0.857±0.023 | 0.843±0.013 |
| Gaussian NB<sup>4</sup> | 0.854±0.064 | 0.639±0.064 | 0.378±0.139 | 0.931±0.023 | 0.858±0.015 | 0.577±0.053 |
| QDA<sup>4</sup> | 0.768±0.059 | 0.699±0.059 | 0.614±0.066 | 0.795±0.075 | 0.773±0.073 | 0.648±0.050 |
| CNN<sup>2</sup> | 0.911±0.006 | 0.827±0.006 | 0.840±0.044 | 0.811±0.051 | 0.836±0.033 | 0.822±0.033 |

<sup>1</sup> Values are mean ± SEM across five folds. Missing values were imputed using $k$ -nearest-neighbor imputation, and predictors were scaled to $[0, 1]$ . <sup>2</sup> CNN, convolutional neural network. Two one-dimensional convolutional layers with 256 and 64 filters, each followed by average pooling, were connected to a 32-unit rectified-linear dense layer and a one-unit sigmoid output. <sup>3</sup> LR, logistic regression; SAGA solver, elastic-net penalty, $\ell_1$ ratio 0.5, and tolerance 0.02. <sup>4</sup> Non-CNN algorithms were implemented using scikit-learn. Default hyperparameters were used unless otherwise shown. <sup>5</sup> AUROC, area under the receiver operating characteristic curve. <sup>6</sup> PPV, positive predictive value. <sup>7</sup> NPV, negative predictive value.

Operating characteristics also differed across algorithms. Gaussian naïve Bayes had high specificity (0.931) but low sensitivity (0.378), whereas AdaBoost and random forest showed more balanced sensitivity and specificity. No algorithm had the highest value for every metric. The combination of broadly similar AUROC values among several algorithms and differences in their operating characteristics supported using the normalized mean-AUROC values as weights in the ensemble attribution analysis.

### 4.2 Variation in Feature Attribution Across Trials and Algorithms

Figure 1 shows patient-level SHAP explanations for four illustrative patients across random forest, Gaussian naïve Bayes, and logistic regression. For the same patient, the algorithms differed in the features receiving the largest attributions and in their attribution magnitudes. Because these patients and algorithms were selected for illustration, the figure does not provide a quantitative estimate of agreement.

**Figure 1:**
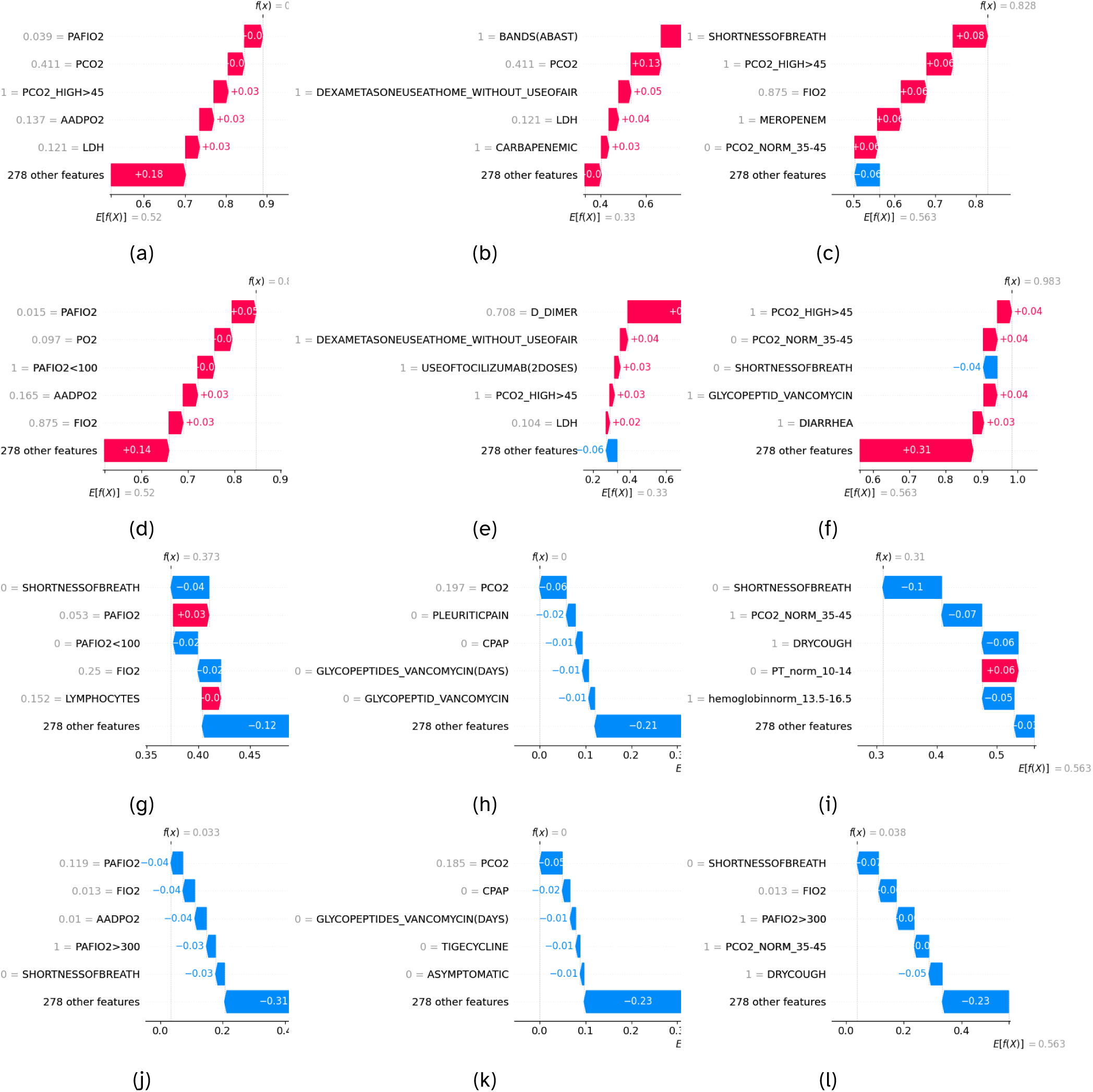
Patient-level SHAP explanations for four illustrative patients across three predictive algorithms. Each row corresponds to one patient: the first two died during hospitalization and the latter two survived to discharge. Columns show random forest, Gaussian naïve Bayes, and logistic regression, respectively. Positive SHAP values increase the explained model output relative to its baseline, whereas negative values decrease it.

Quantitative comparison of the normalized global importance profiles supported the pattern illustrated in Figure 1. Across 1,350 unique comparisons, repeated trials of the same algorithm had a mean Pearson correlation of 0.783 *±* 0.004 SEM. Across 1,350 comparisons of different algorithms fitted within the same trial, the mean correlation was 0.369 *±* 0.006 SEM (Table 2). Thus, profiles were more similar across changes in training samples within an algorithm than across algorithm choices, although both sources contributed to attribution variation.

**Table 2:** Similarity of normalized SHAP feature-importance profiles across training trials and predictive algorithms.

| Comparison | Number of pairs | Pearson $r$, mean ± SEM |
| --- | --- | --- |
| Same algorithm across training trials | 1,350 | $0.783 \pm 0.004$ |
| Different algorithms within a trial | 1,350 | $0.369 \pm 0.006$ |

### 4.3 Normalized Ensemble Feature Importance and Attribution Alignment

Table 3 reports two complementary summaries for 250 non-vaccination features. Importance is the mean absolute normalized ensemble attribution across 30 trials, with trial-based 95% confidence intervals. Alignment is the mean cosine similarity between signed attribution vectors for corresponding patients across unique trial pairs; the accompanying *q*-value comes from the patient-correspondence randomization analysis after false-discovery-rate adjustment. Importance therefore measures average magnitude, whereas alignment measures the consistency of the patient-level attribution pattern.

**Table 3:**
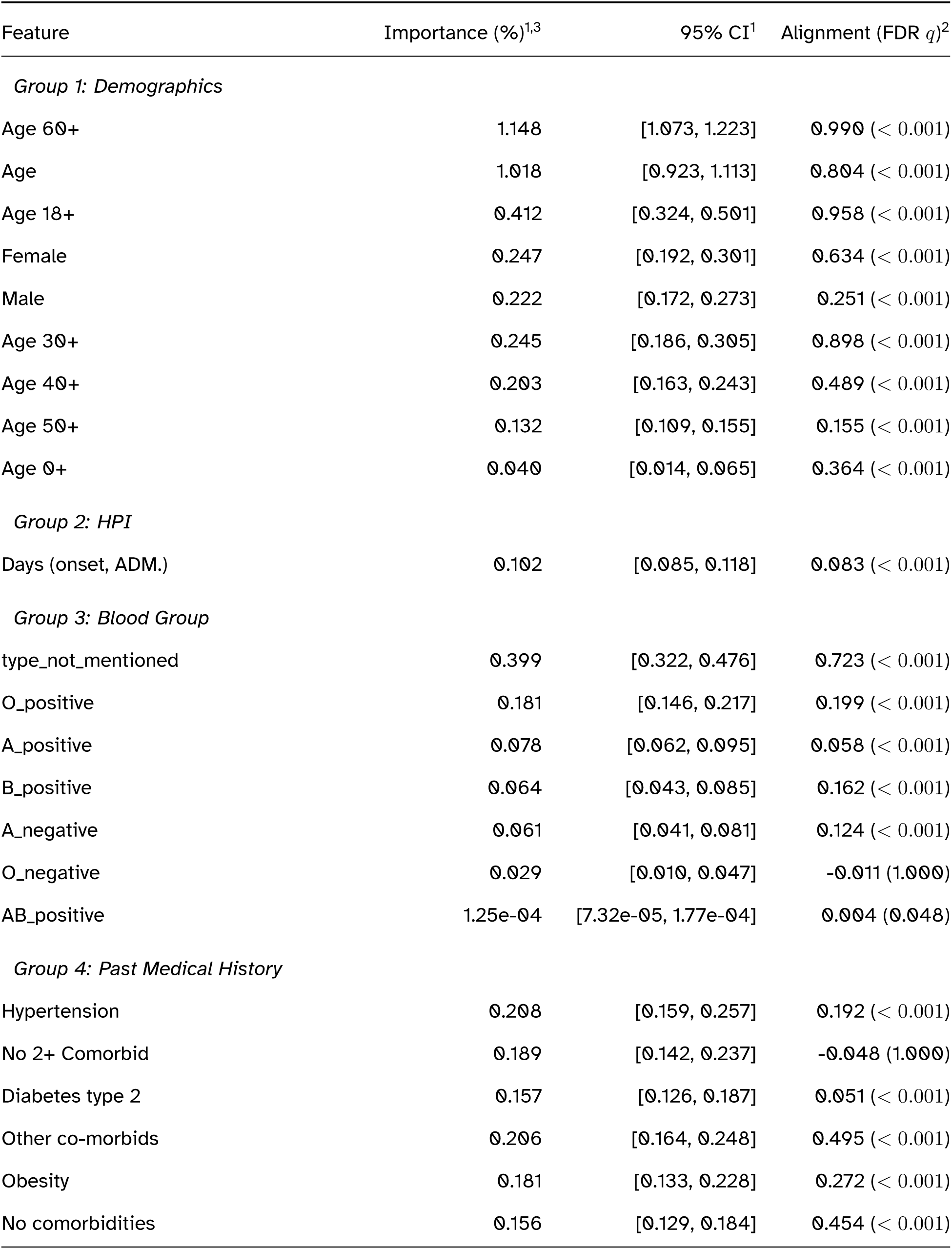
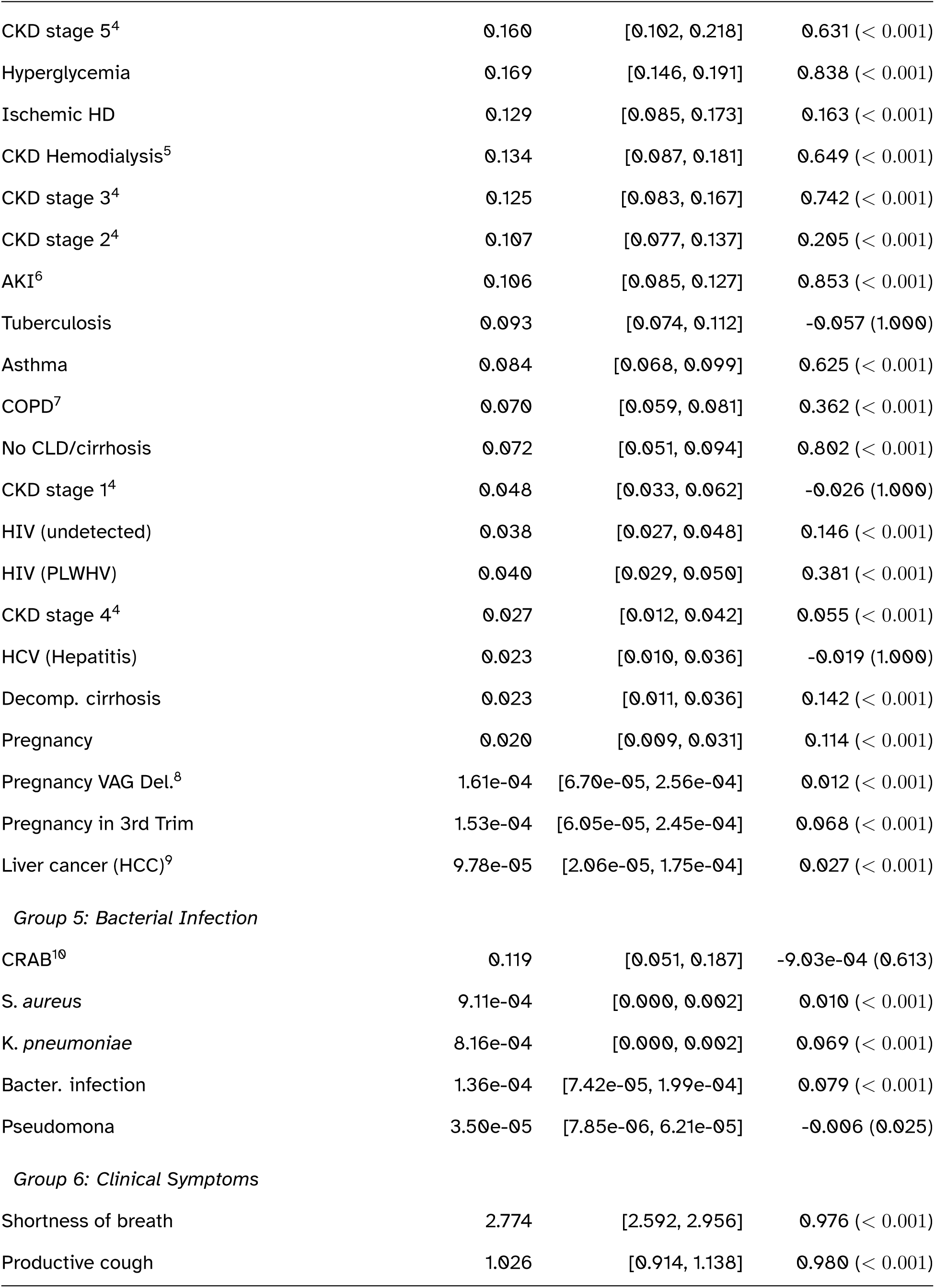
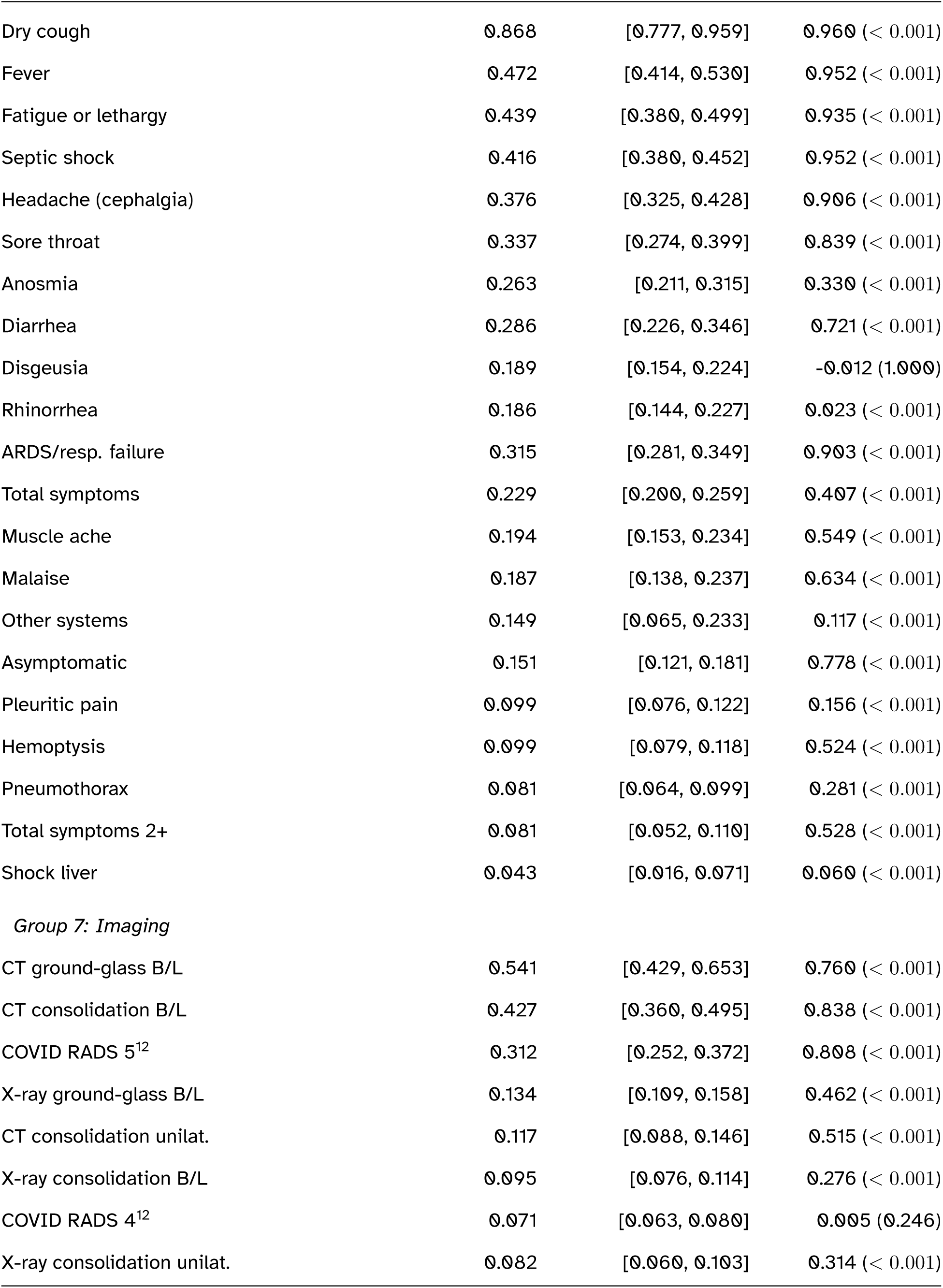
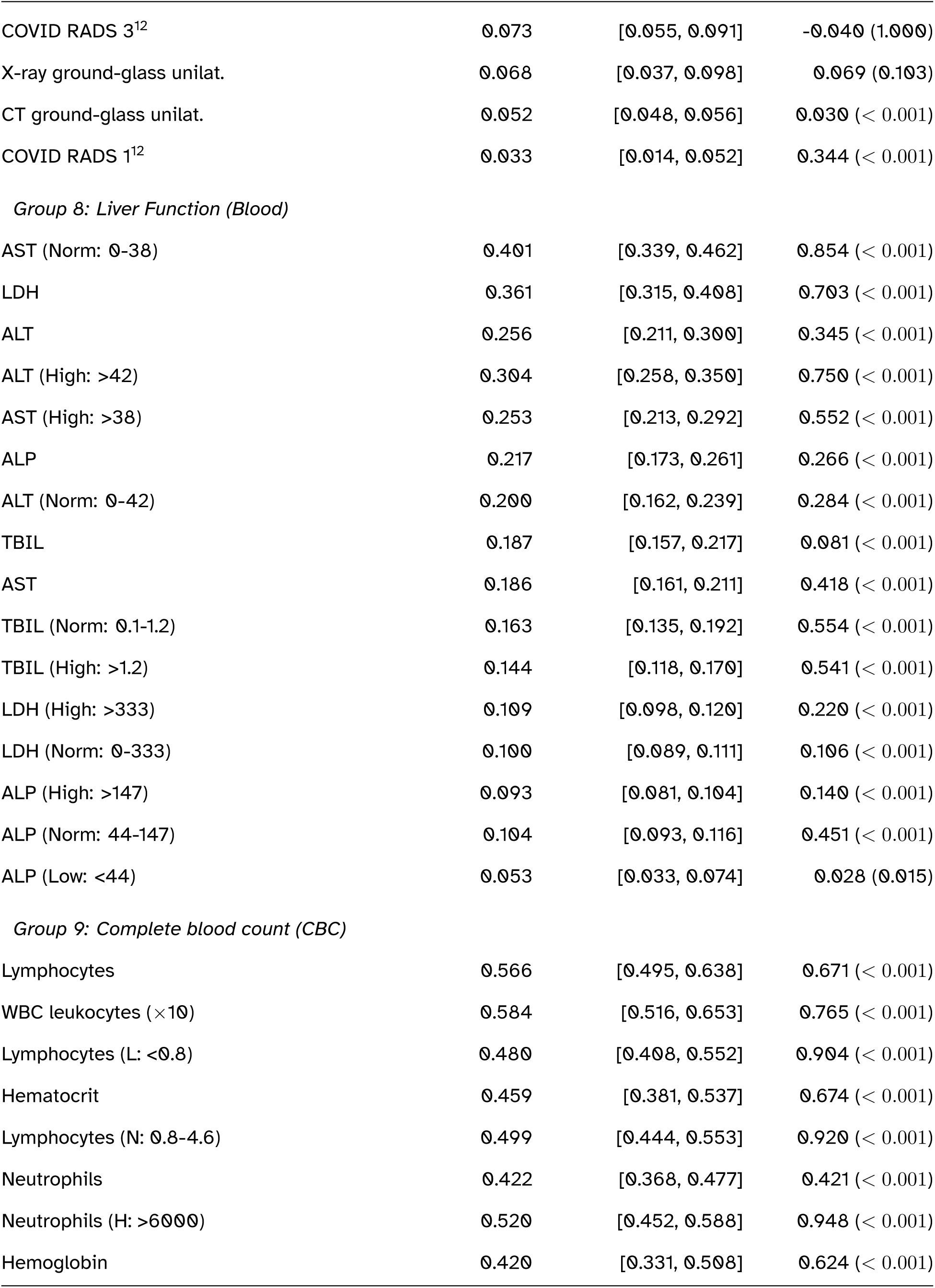
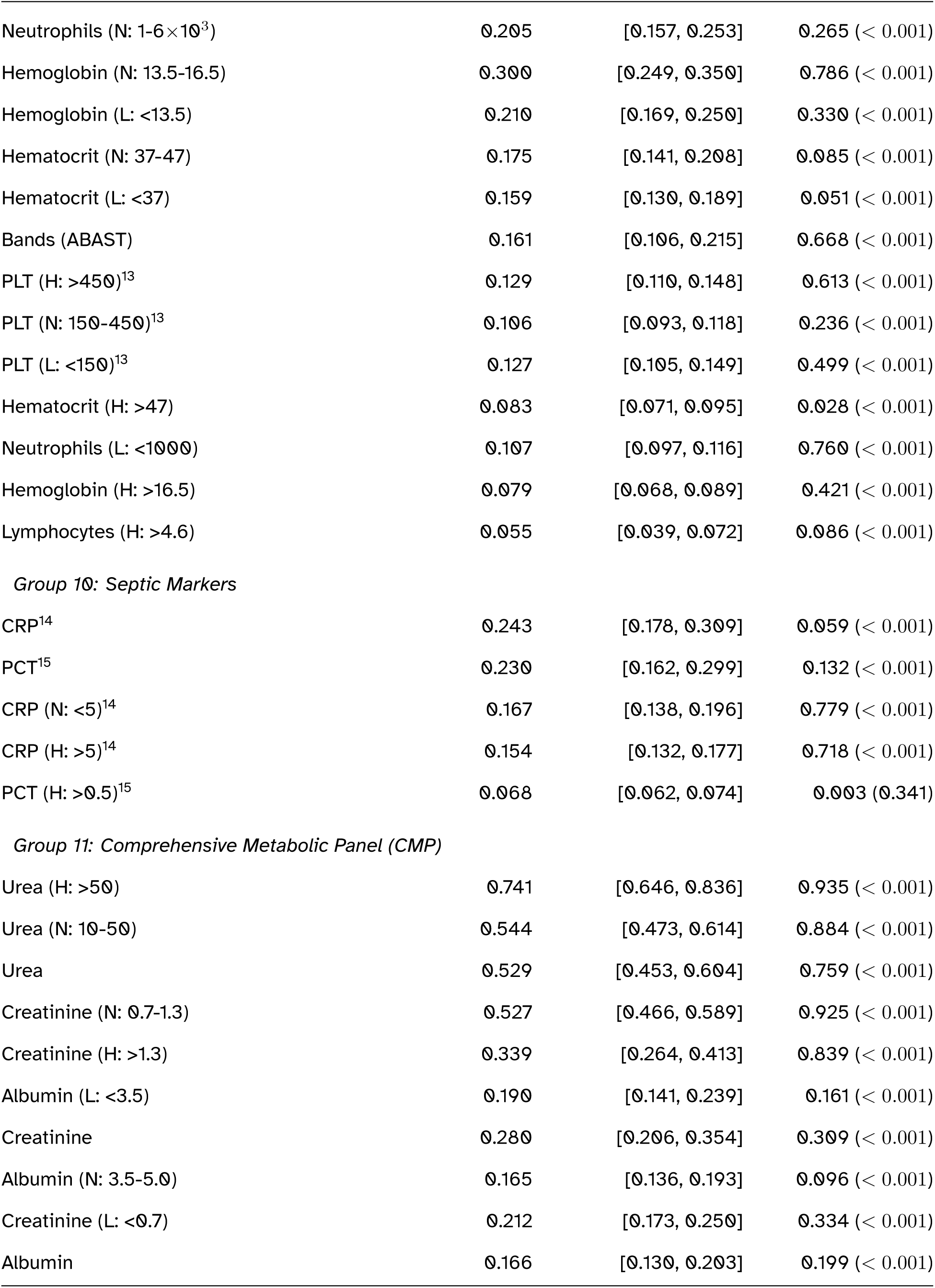
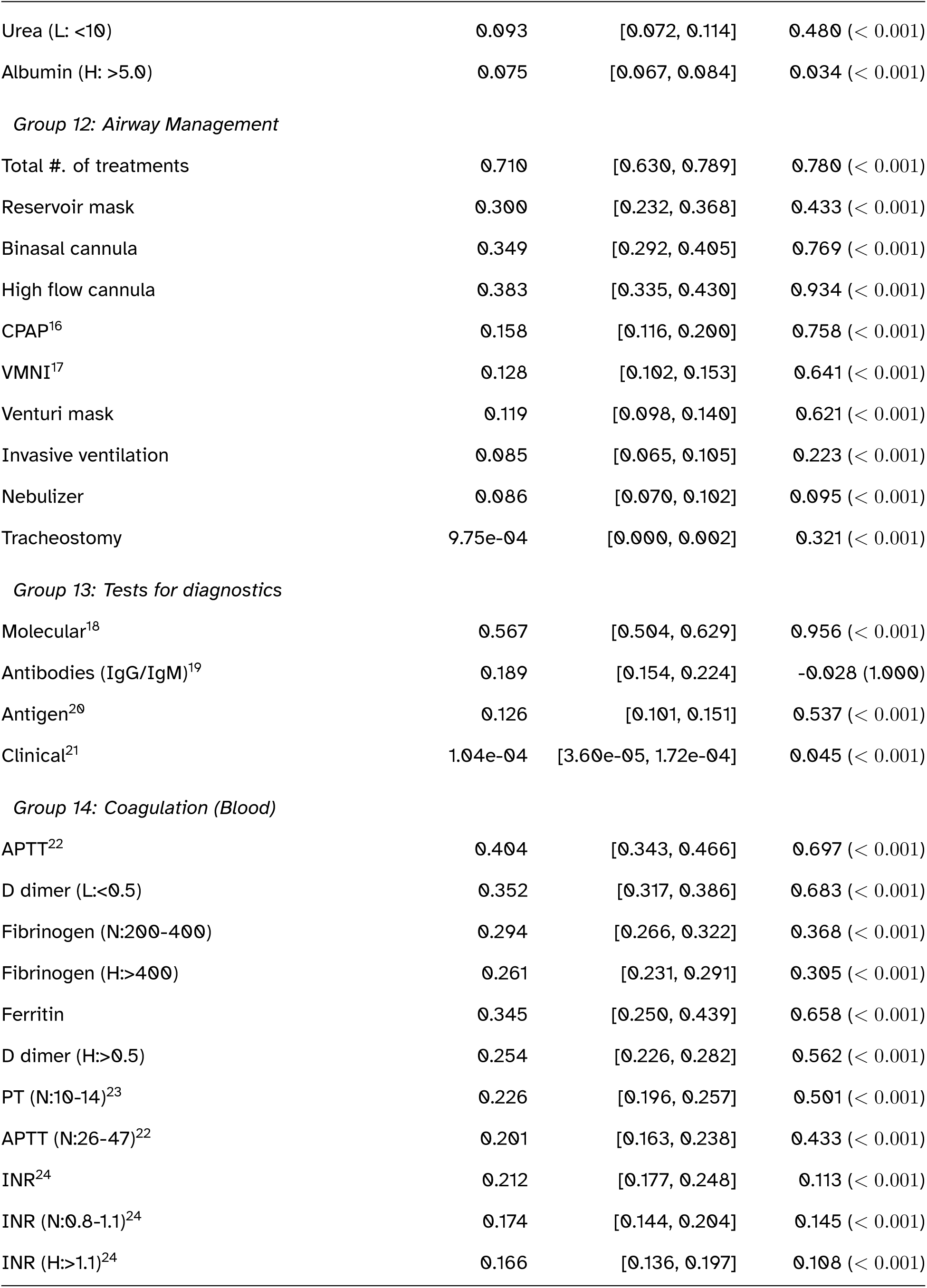
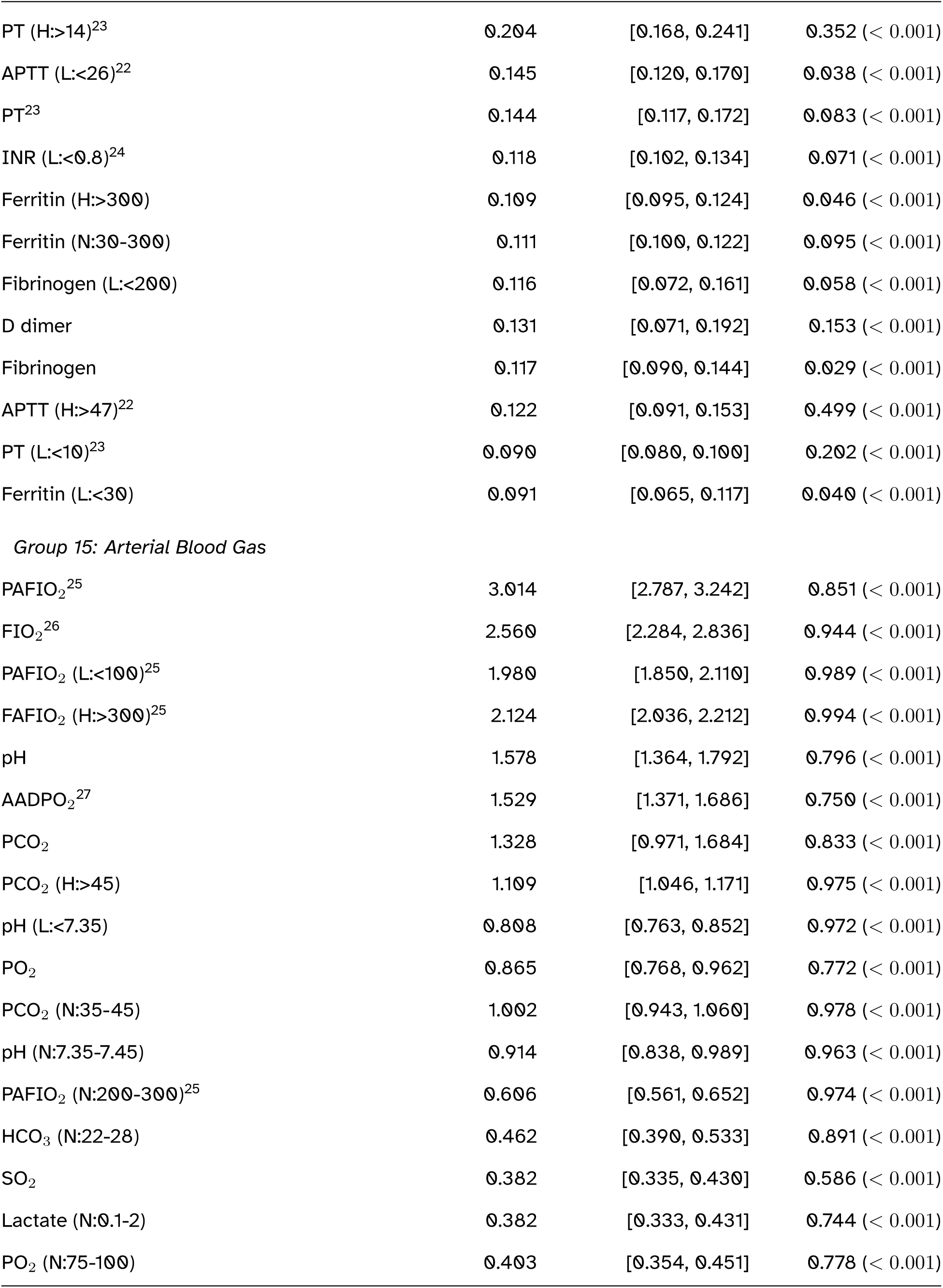
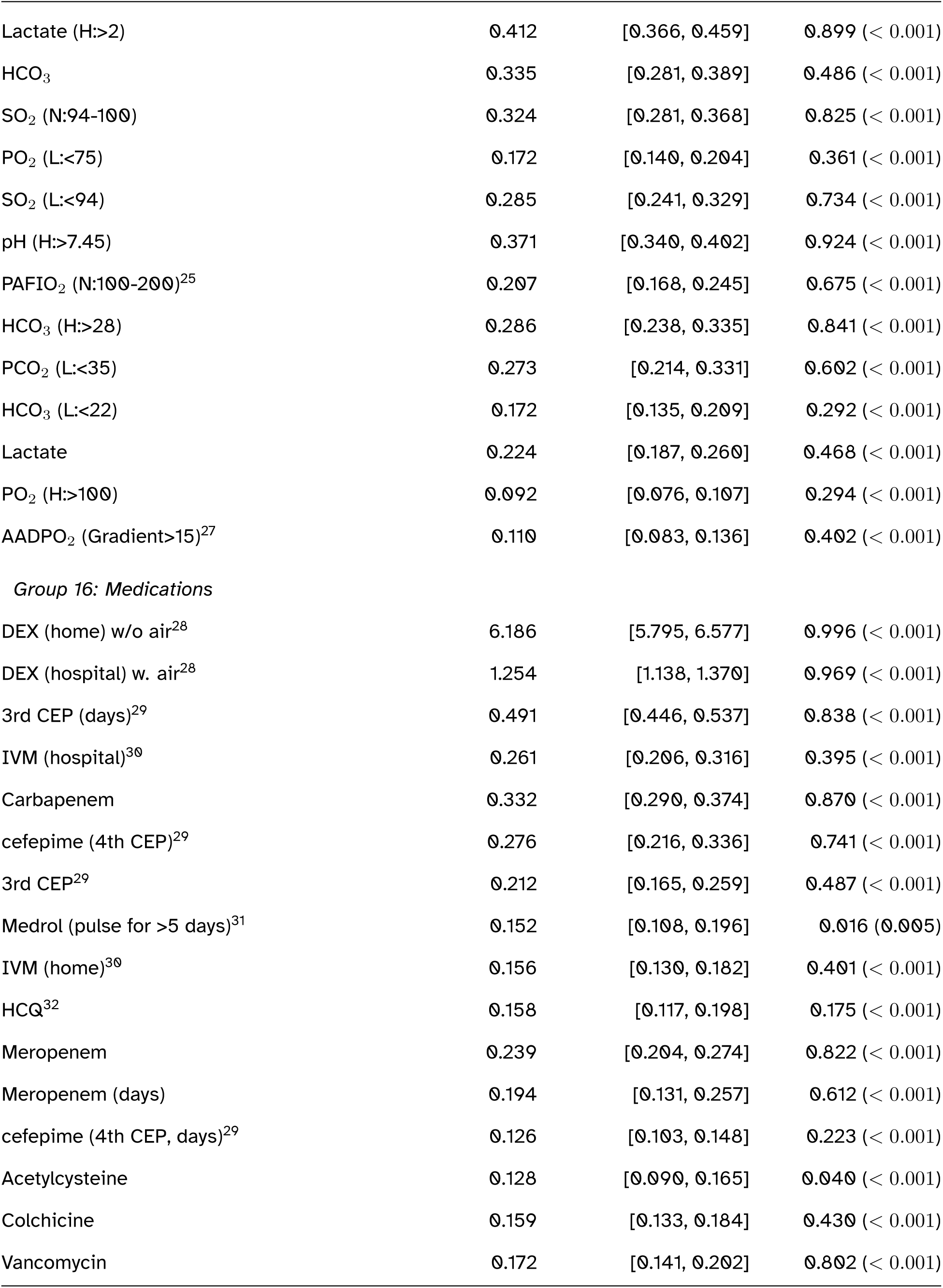
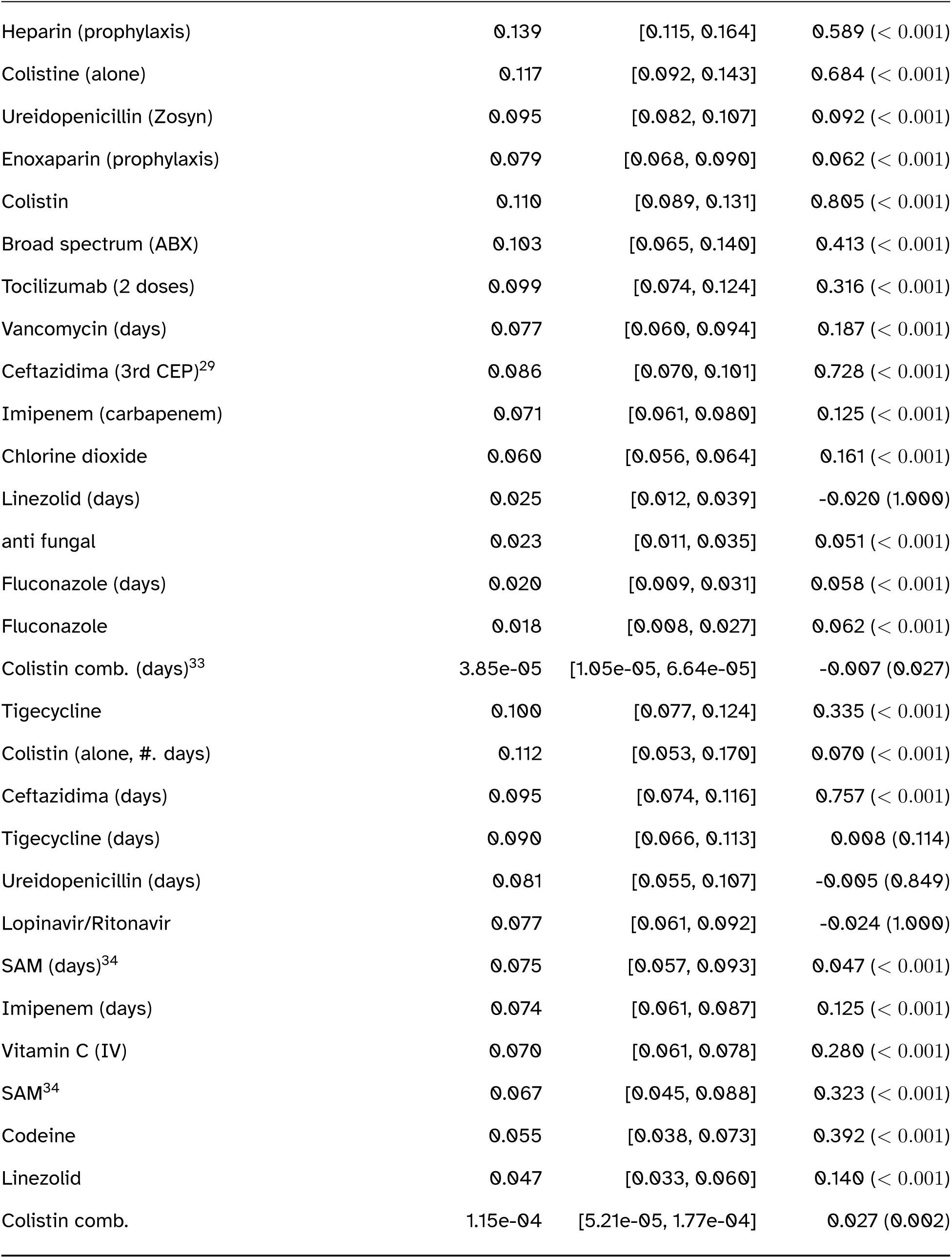
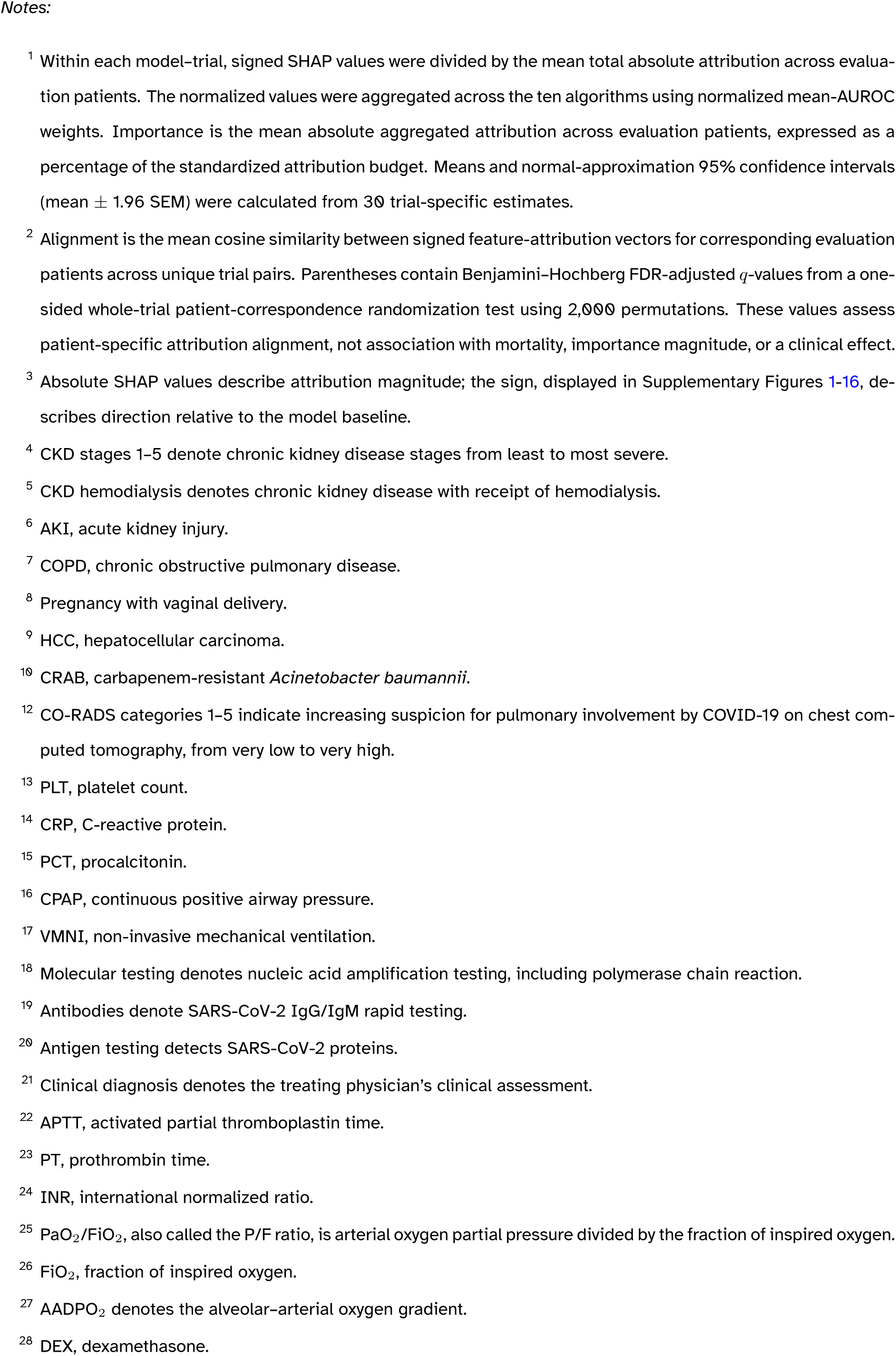
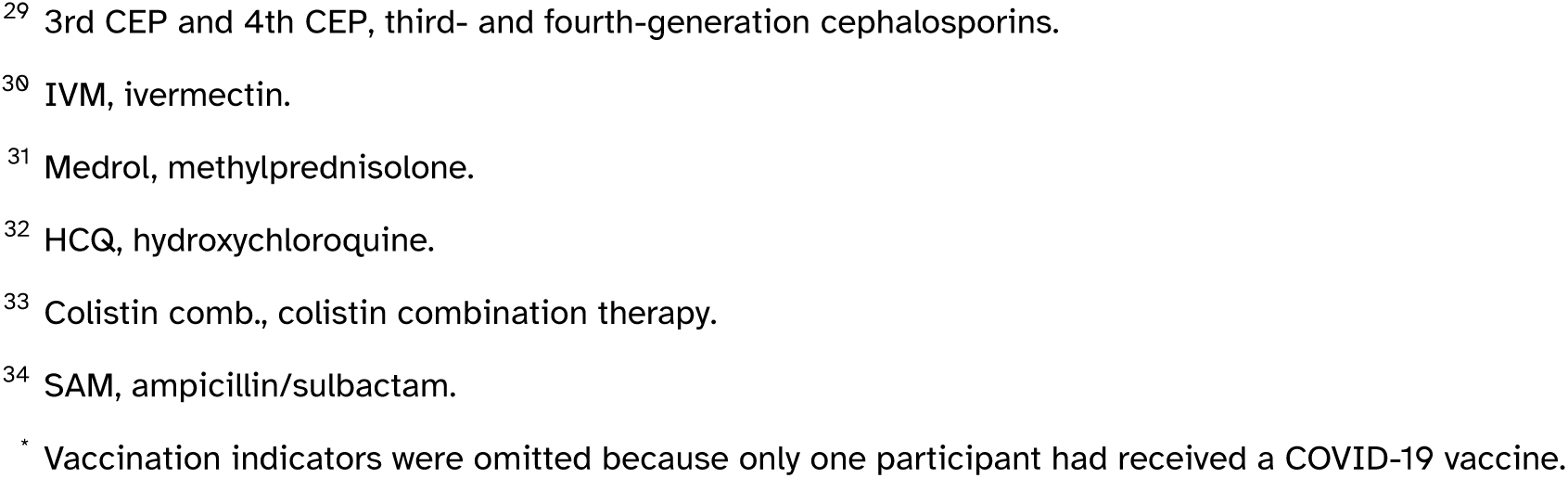
Normalized ensemble SHAP feature importance and patient-level attribution alignment across 30 resampled training trials, organized into 16 clinical categories.

The largest normalized ensemble importance was observed for dexamethasone use at home without oxygen support (6.186%; 95% CI, 5.795–6.577), followed by the continuous *PaO*_2_/*FiO*_2_ ratio (3.014%; 95% CI, 2.787–3.242), shortness of breath (2.774%; 95% CI, 2.592–2.956), and *FiO*_2_ (2.560%; 95% CI, 2.284–2.836). These features also had high mean patient-level alignment, ranging from 0.851 for *PaO*_2_/*FiO*_2_ to 0.996 for home dexamethasone use, with *q <* 0.001 for each.

#### 4.3.1 Demographic and Baseline Health Indicators

The indicator for age 60 years or older had an importance of 1.148% (95% CI, 1.073–1.223), and continuous age had an importance of 1.018% (95% CI, 0.923–1.113). Their mean alignments were 0.990 and 0.804, respectively (*q <* 0.001 for both). The sex indicators had lower importance: 0.247% for female and 0.222% for male. Among past medical history variables, other recorded comorbidities (0.206%), hypertension (0.208%), obesity (0.181%), hyperglycemia (0.169%), stage 5 chronic kidney disease (0.160%), and type 2 diabetes (0.157%) contributed to the ensemble profile at broadly similar magnitudes.

Importance magnitude did not uniformly imply patient-level alignment. For example, the indicator for fewer than two comorbidities had an importance of 0.189% but a mean alignment of *−*0.048 (*q* = 1.000), whereas hyperglycemia had a similar importance of 0.169% and an alignment of 0.838 (*q <* 0.001). This difference illustrates the distinct information provided by the two columns of Table 3.

#### 4.3.2 Clinical Presentation and Symptoms

Shortness of breath was the leading symptom feature (2.774%; 95% CI, 2.592–2.956), followed by productive cough (1.026%), dry cough (0.868%), fever (0.472%), fatigue or lethargy (0.439%), and septic shock (0.416%). These features showed high mean patient-level alignment, ranging from 0.935 to 0.980, with *q <* 0.001 for each. Dysgeusia had a lower importance of 0.189% and did not show corresponding alignment (mean cosine similarity, *−*0.012; *q* = 1.000).

#### 4.3.3 Laboratory Measurements

Laboratory contributions spanned hematologic, renal, hepatic, inflammatory, coagulation, and blood-gas measurements. Elevated urea had an importance of 0.741%, while continuous urea and normal-range urea had values of 0.529% and 0.544%, respectively. Normal-range creatinine had an importance of 0.527%. Among complete blood count variables, leukocyte count and lymphocyte count had importance values of 0.584% and 0.566%. Other contributions included APTT (0.404%), normal-range AST (0.401%), LDH (0.361%), low D-dimer (0.352%), ferritin (0.345%), C-reactive protein (0.243%), and procalcitonin (0.230%). These values identify information used by the fitted models and do not establish organ injury or a temporal sequence of physiologic deterioration.

#### 4.3.4 Imaging and Physiologic Measurements

Oxygenation and arterial blood-gas variables contained several of the largest attribution magnitudes. In addition to continuous *PaO*_2_/*FiO*_2_ and *FiO*_2_, *PaO*_2_/*FiO*_2_ *<* 100 had an importance of 1.980%, pH 1.578%, the alveolar–arterial oxygen gradient 1.529%, and *PCO*_2_ 1.328%. Bilateral ground-glass opacity and bilateral consolidation on computed tomography had importance values of 0.541% and 0.427%, respectively. These results show that physiologic and imaging measurements both contributed to the mortality predictions, with larger attribution magnitudes for several oxygenation measures.

#### 4.3.5 Acute Care Interventions and Pharmacotherapy

Treatment and respiratory-support variables also contributed to the ensemble profile. In addition to home dexamethasone use without oxygen support, dexamethasone administered in hospital with oxygen support had an importance of 1.254%, total number of treatments 0.710%, third-generation cephalosporin duration 0.491%, high-flow cannula 0.383%, and binasal cannula 0.349%. These attributions describe patterns captured by the models and may reflect disease severity, treatment indication, timing, access to care, and treatment selection. They do not estimate treatment benefit or harm.

### 4.4 Distribution-Adjusted Subgroup Differences

Table 4 presents, within each subgroup, up to 20 features with the largest absolute distribution-adjusted differences in normalized ensemble SHAP importance. The pooled evaluation reference for every comparison comprised all 558 available evaluation observations, including observations belonging to the subgroup. Differences were calculated as pooled-reference importance minus distribution-adjusted subgroup importance. Positive values therefore indicate greater attribution magnitude in the pooled evaluation reference, whereas negative values indicate greater magnitude in the distribution-adjusted subgroup. All 283 encoded features entered the calculations; rows were selected by *|*Δ*|*, rather than by the nominal *p*-value.

**Table 4:**
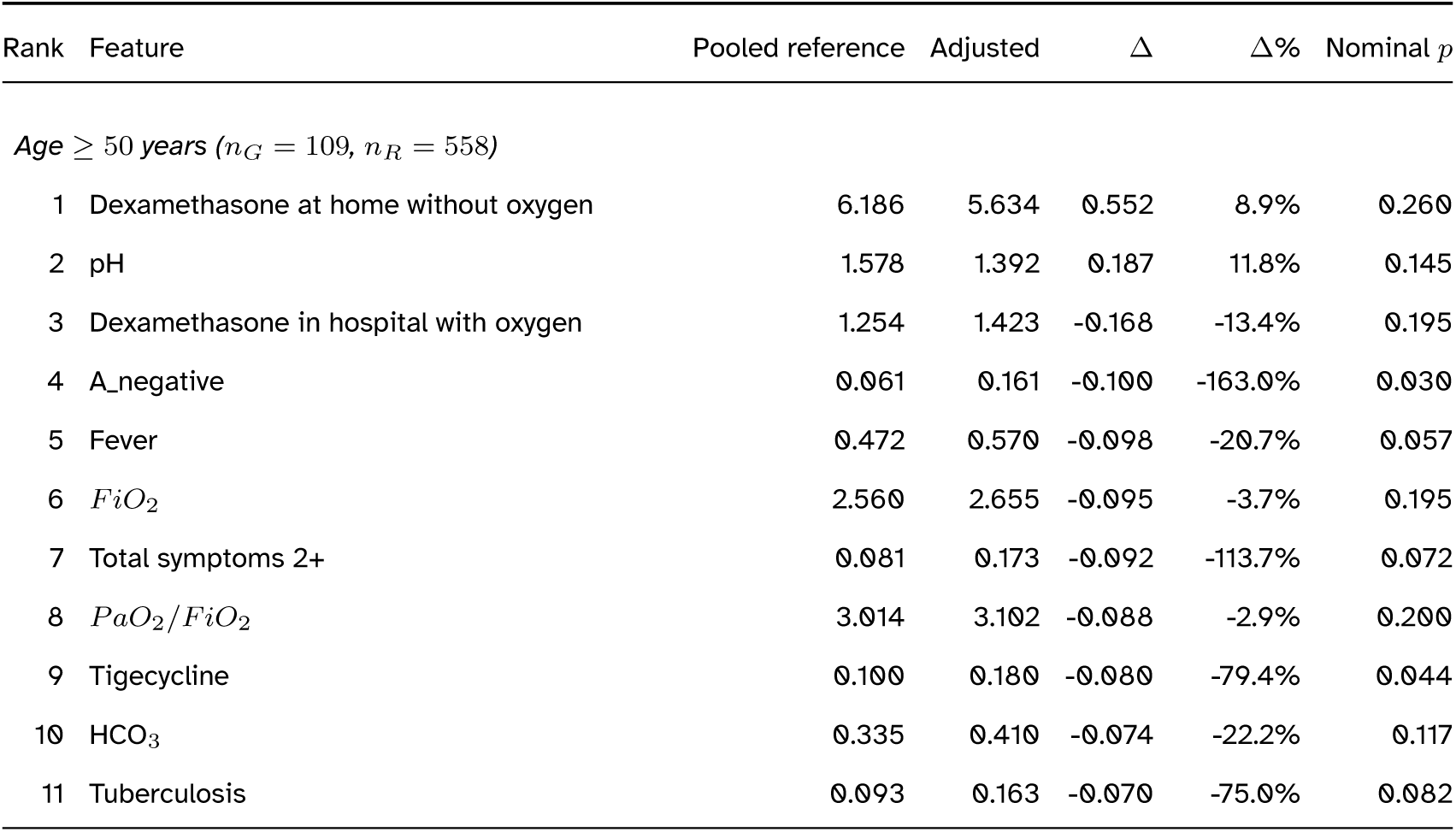
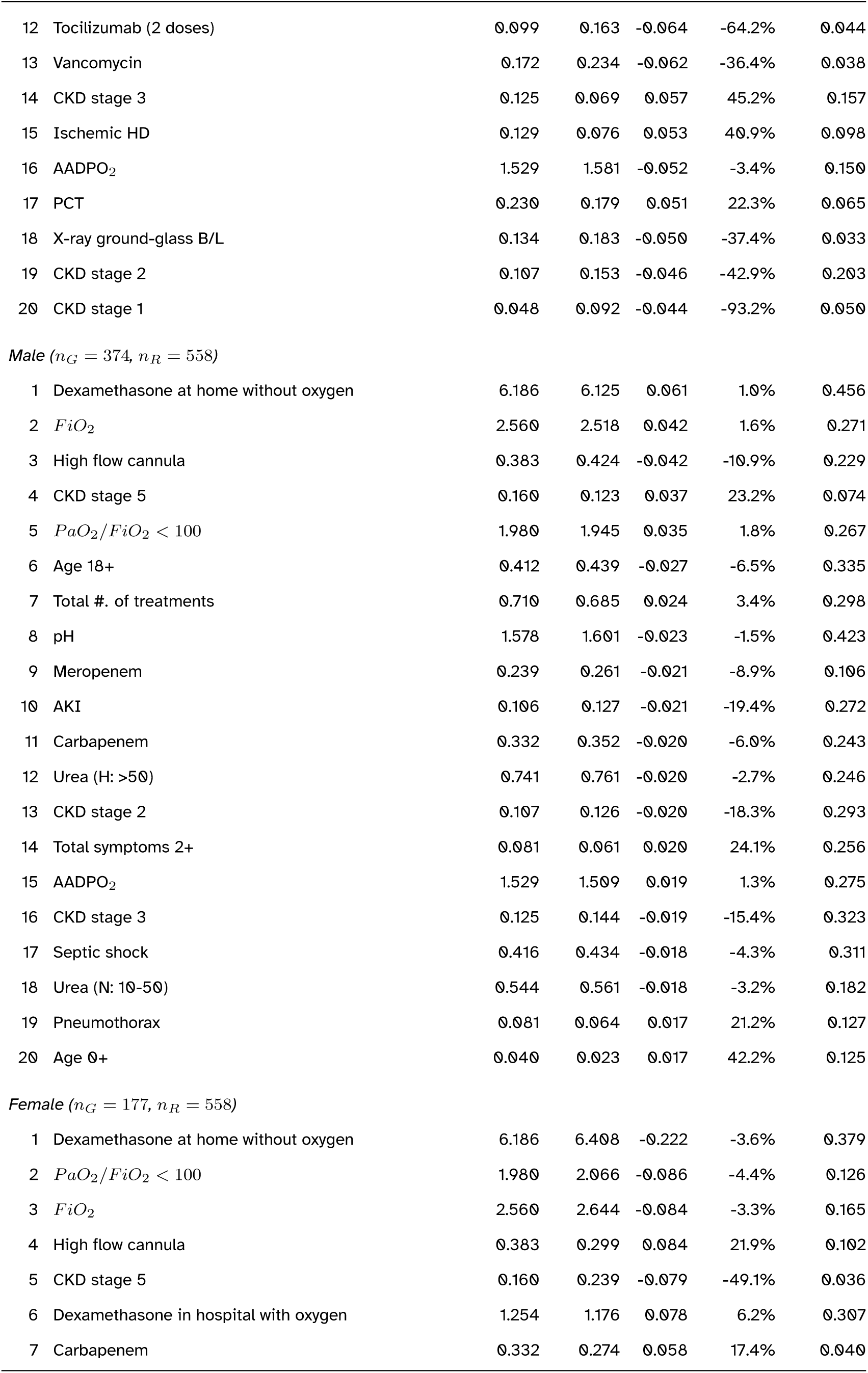
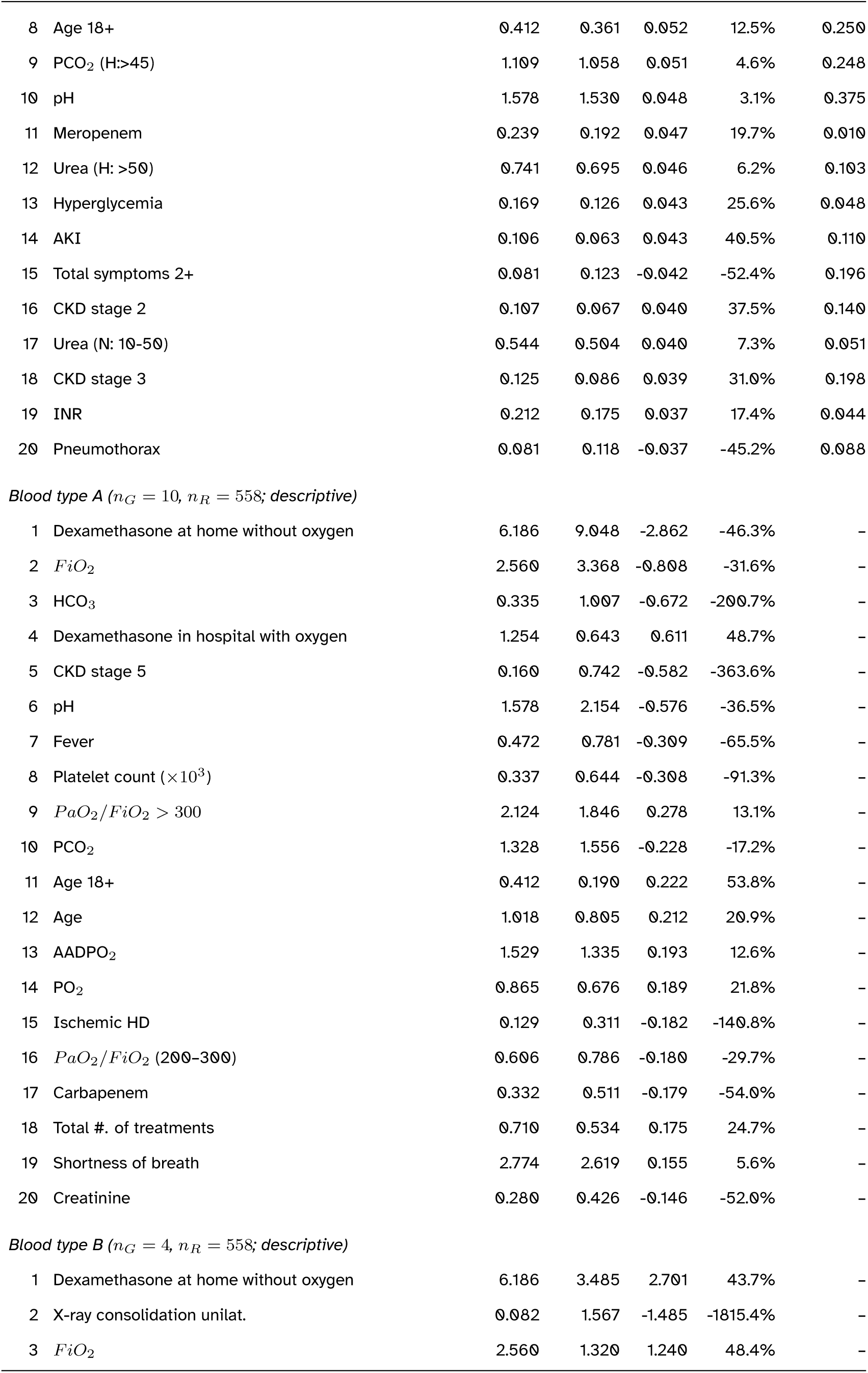
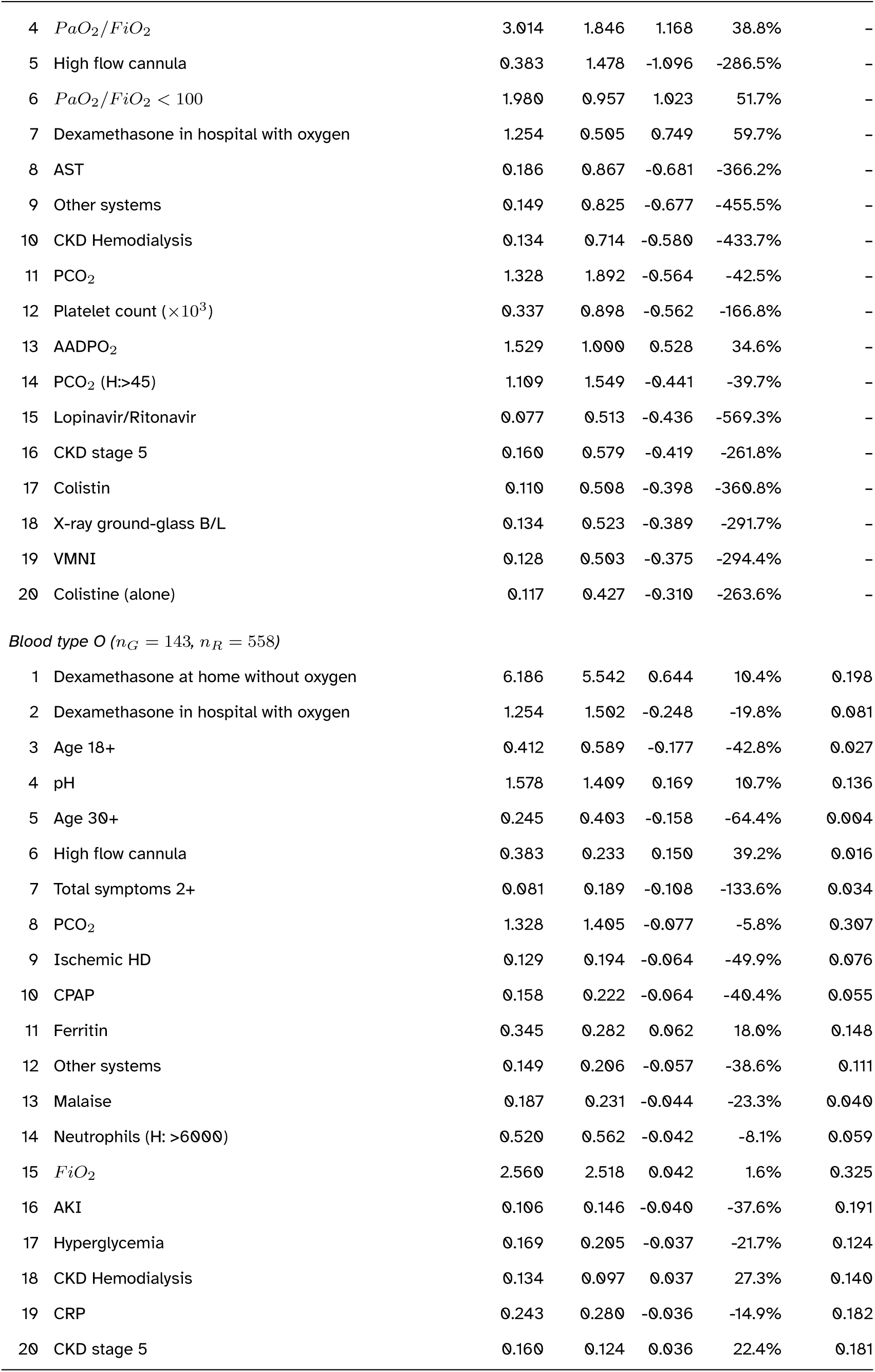
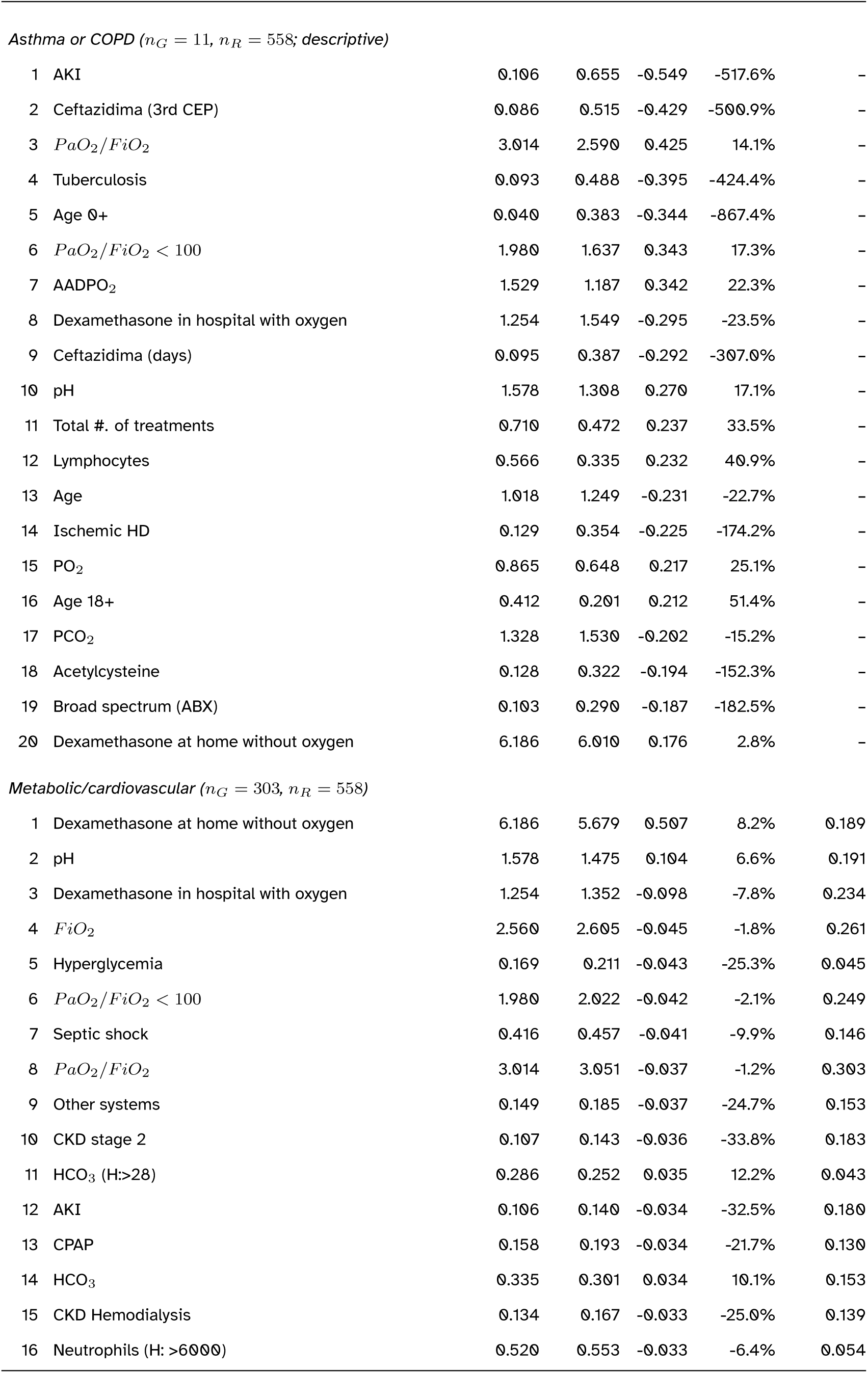
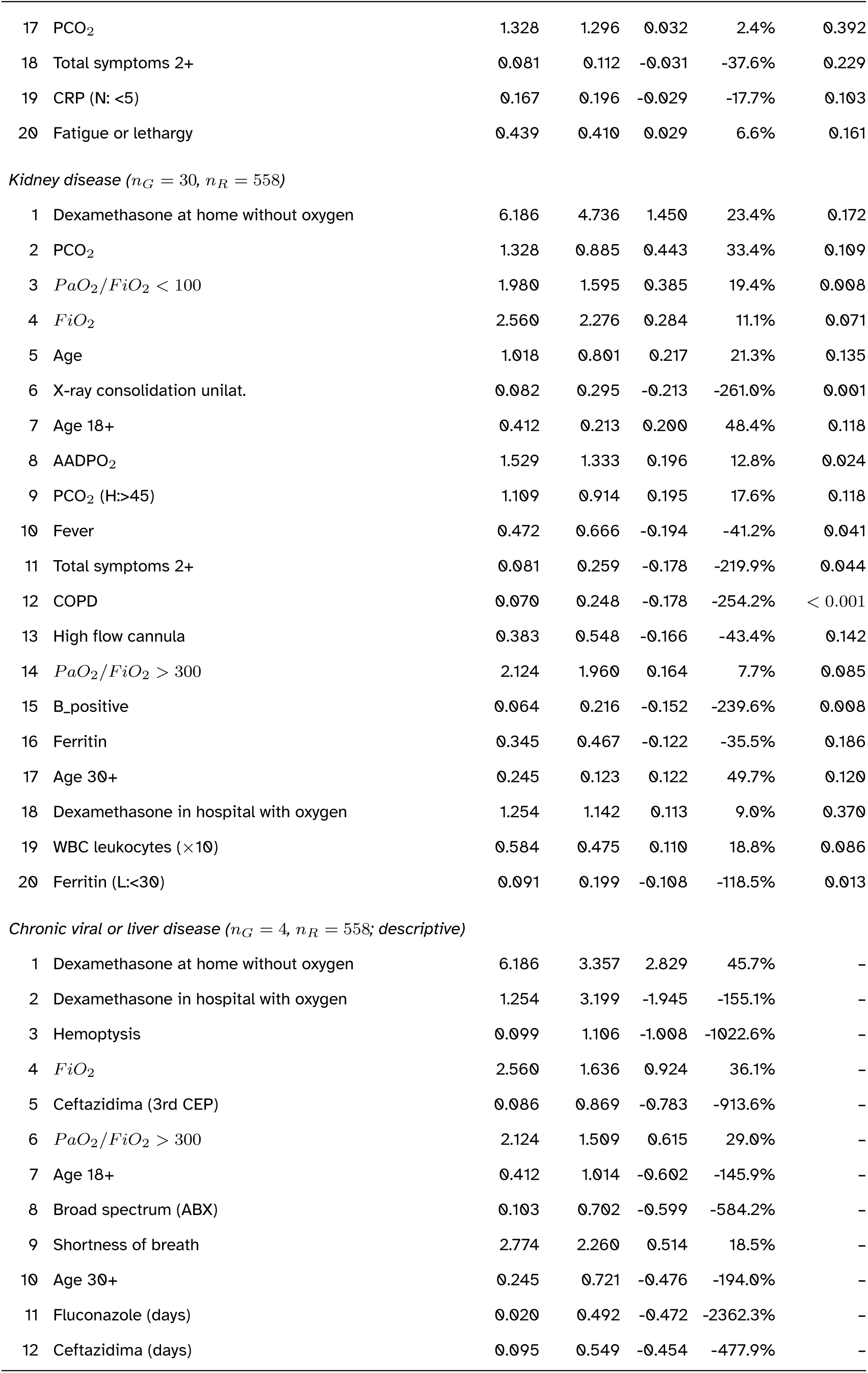
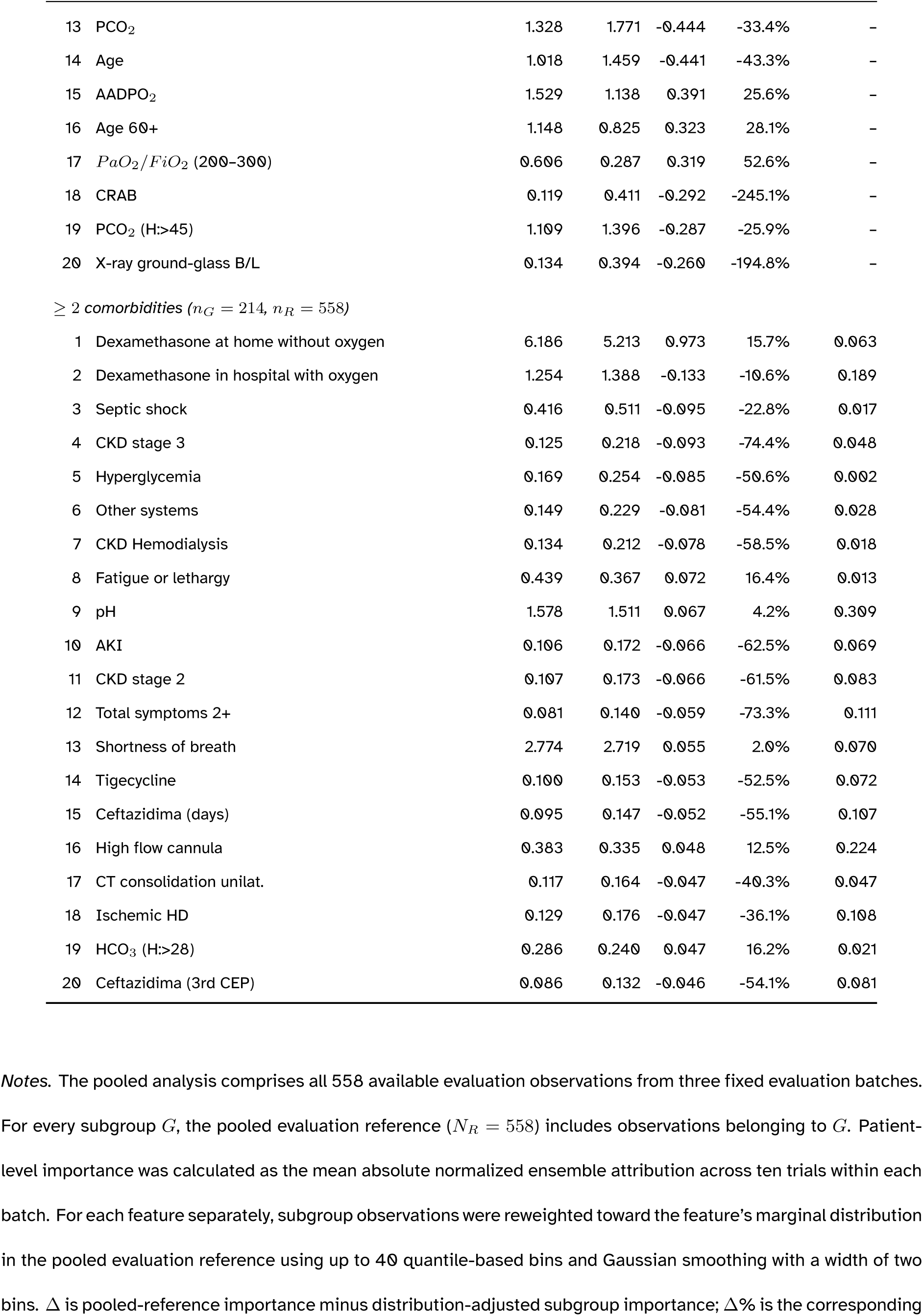
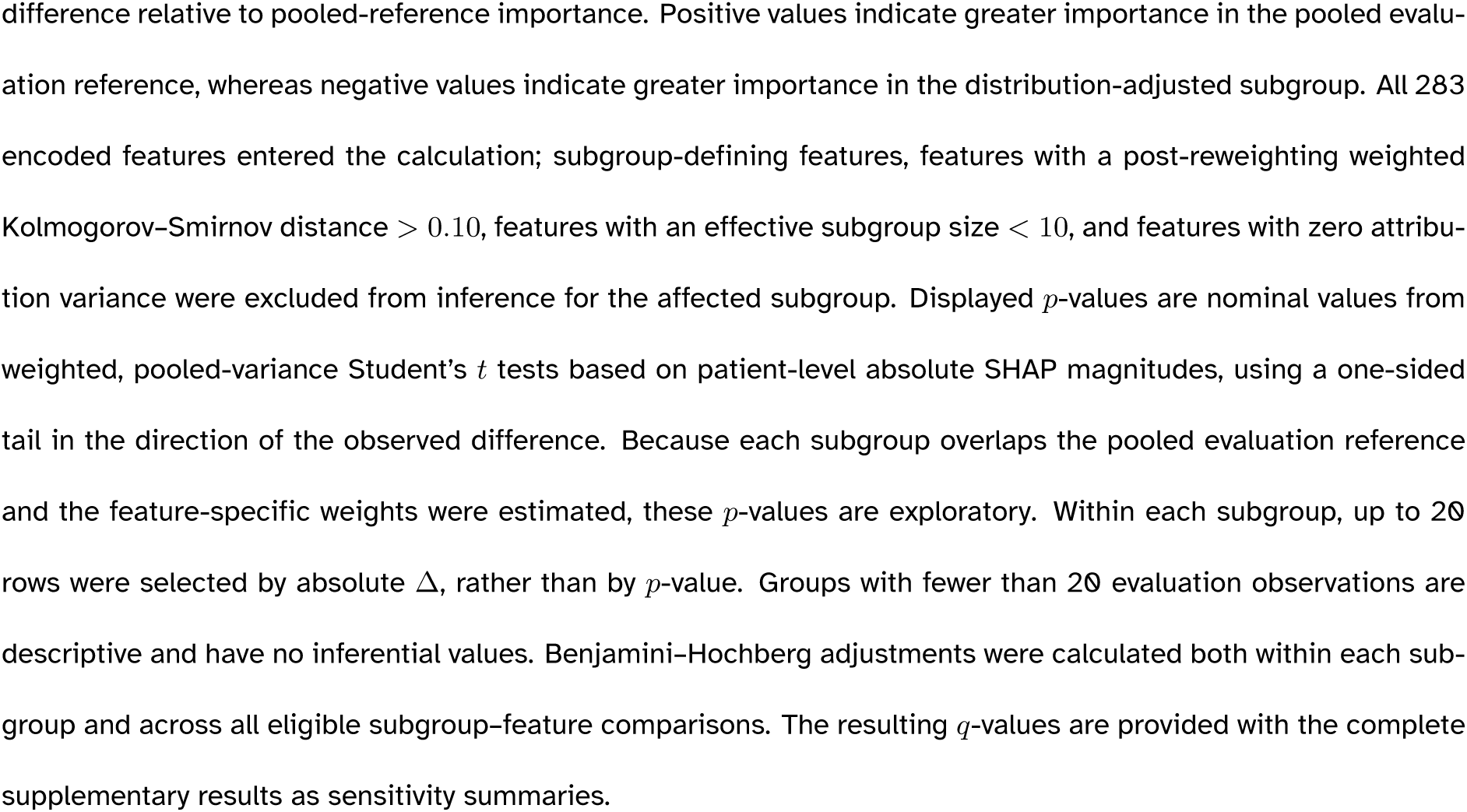
The largest absolute distribution-adjusted differences in normalized ensemble SHAP importance between each subgroup and the pooled evaluation reference. Within each subgroup, up to 20 features were selected by absolute Δ, rather than by nominal *p*-value.

Several of the largest absolute differences occurred in the kidney-disease subgroup. Home dexamethasone use without oxygen support had lower adjusted-subgroup importance than pooled-reference importance (4.736% versus 6.186%; Δ = 1.450; 23.4%), although its nominal one-sided *p*-value was 0.172. Adjusted importance for *PaO*_2_/*FiO*_2_ *<* 100 was also lower than pooled-reference importance in this subgroup (1.595% versus 1.980%; Δ = 0.385; 19.4%; nominal *p* = 0.008).

Among evaluation observations from patients with at least two documented comorbidities, adjusted importance was lower than pooled-reference importance for home dexamethasone use without oxygen support (Δ = 0.973; 15.7%; nominal *p* = 0.063) and higher for septic shock (Δ = *−*0.095; *−*22.8%; nominal *p* = 0.017) and hyperglycemia (Δ = *−*0.085; *−*50.6%; nominal *p* = 0.002). In the type O subgroup, age 30 years or older had greater adjusted importance than in the pooled evaluation reference (Δ = *−*0.158; *−*64.4%; nominal *p* = 0.004). Results for blood types A and B, asthma or COPD, and chronic viral or liver disease were descriptive because these subgroups contained fewer than 20 evaluation observations.

Across 1,763 eligible subgroup–feature comparisons, 64 had nominal one-sided *p <* 0.05. None remained below 0.05 after Benjamini–Hochberg adjustment within each subgroup or across all comparisons. Because these tests do not fully account for subgroup–reference overlap or uncertainty in the estimated reweighting weights, the findings are exploratory and do not establish clinical mechanisms or treatment effects.

## 5 Discussion

### 5.1 Principal Findings

Algorithms with broadly similar predictive performance produced different feature-attribution profiles. Mean correlations were higher across repeated training trials of the same algorithm (*r* = 0.783) than across different algorithms within the same trial (*r* = 0.369), indicating that model choice contributed more strongly to global attribution variation than changes in the sampled training set. Performance-weighted ensemble SHAP aggregation provided a representative relative-importance profile across the included algorithms, while trial-based confidence intervals and patient-level alignment characterized complementary aspects of attribution variability.

The aggregated profile emphasized oxygenation measures, blood-gas variables, symptoms, renal and metabolic indicators, imaging findings, and treatment-related variables. Distribution-adjusted subgroup comparisons provided an additional, exploratory view of how attribution magnitude varied across patient groups after adjusting each feature’s marginal distribution toward the pooled evaluation reference. Of 1,763 eligible subgroup–feature comparisons, 64 had nominal one-sided *p <* 0.05, but none remained below 0.05 after Benjamini–Hochberg adjustment. The subgroup results should therefore be interpreted as candidate patterns for further study rather than evidence of established subgroup-specific effects.

### 5.2 Clinical Interpretation of the Attribution Profile

Oxygenation and blood-gas measures appeared alongside renal, metabolic, and coagulation indicators, suggesting that the fitted models drew on information from several physiologic domains. This pattern is compatible with the pulmonary and extrapulmonary manifestations described in COVID-19 [6]. The prominence of oxygenation and urea measurements also parallels their inclusion in the validated 4C Mortality Score [10], while the contribution of coagulation variables is consistent with reports linking coagulation abnormalities to adverse outcomes [19]. These comparisons support the clinical plausibility of the attribution profile but do not establish a temporal sequence of organ dysfunction or identify causal pathways.

Shortness of breath and cough received substantial attribution alongside blood-gas and laboratory measurements. Clinical cohort studies have similarly considered presenting symptoms together with physiologic and laboratory findings when characterizing COVID-19 outcomes [25]. Their joint prominence in our analysis suggests that clinical presentation and objective measurements provided complementary predictive information. The analysis did not, however, quantify the independent predictive gain attributable to each feature group.

Treatment-related variables, including dexamethasone use and respiratory support, require contextual interpretation because their attributions may reflect disease severity, treatment selection, timing, and care setting. In the RECOVERY trial, dexamethasone reduced mortality among patients receiving oxygen or invasive mechanical ventilation, whereas no mortality benefit was demonstrated among patients receiving no respiratory support [20]. This evidence emphasizes the importance of treatment context; it does not validate the direction or magnitude of treatment attributions in our cohort. The present findings describe treatment and care patterns captured by the models and do not estimate treatment benefit or harm.

### 5.3 Subgroup Patterns and Hypotheses

The subgroup comparisons suggest that the relative predictive information contributed by individual features may vary with patient characteristics. Differences involving oxygenation, symptoms, renal measures, and metabolic indicators motivate the hypothesis that baseline health and physiologic reserve influence which measurements are most informative during hospitalization. However, none of the subgroup–feature comparisons remained below 0.05 after false-discovery-rate adjustment, and several clinically defined subgroups were too small for inferential analysis. These patterns therefore require evaluation in larger independent cohorts with longitudinal measurements and prespecified subgroup hypotheses before they can inform clinical monitoring or treatment.

### 5.4 Relationship to Prior Clinical and Machine Learning Studies

Previous COVID-19 studies used predictive models and feature interpretation to identify informative patient characteristics and examine their contributions to mortality predictions [23, 2, 18]. Our findings similarly emphasize age, oxygenation, symptoms, and laboratory measurements. The present study extends these applications by examining attribution profiles across algorithms and repeated training trials and by comparing subgroup profiles after feature-specific distribution adjustment.

Methodologically, prior work has characterized importance across well-performing models, pooled Shapley-based estimates, and aggregated explanations [4, 16, 1]. Our contribution lies in integrating performance-weighted SHAP aggregation with trial-based importance summaries, patient-level attribution alignment, and distribution-adjusted subgroup comparison in a clinical cohort. This workflow places a representative importance profile alongside information about its variation across fitted models and patient groups, providing interpretive context that is unavailable from a single fitted-model explanation.

### 5.5 Strengths, Limitations, and Future Directions

Strengths include a prospectively collected cohort from two hospitals in Peru, evaluation of ten predictive algorithms, and comparison of feature attributions across repeated training trials. The analysis distinguishes global importance magnitude from the consistency of signed patient-level attribution patterns. Distribution-adjusted subgroup analysis provides a further descriptive view of heterogeneity in model explanations.

Several limitations concern the clinical data and study setting. The cohort was drawn from two hospitals during the early pandemic, and patients with documented psychiatric conditions were excluded, limiting generalizability to other populations, care settings, and pandemic periods. Clinical variables were collected from enrollment throughout hospitalization until discharge or death. Because the analysis did not impose a common prediction time or account for when each variable became available, the reported performance and attributions characterize information observed during the hospital course rather than a deployable admission-time prediction model. External validation with a prespecified prediction time is needed.

The resampling design also limits interpretation of attribution robustness. Training and evaluation subsets were selected independently, so some evaluation observations may also have contributed to model fitting. This overlap may have increased apparent agreement across trials. In addition, original patient identifiers were unavailable across the three evaluation batches; repeated individuals between batches, if present, could therefore not be identified or accounted for. Cross-validated AUROC weights were estimated from the full study cohort, including observations later used in attribution evaluation. The resulting estimates characterize internal model behavior and should not be interpreted as out-of-sample evidence of explanation transportability.

The uncertainty summaries have additional limitations. The normal-approximation 95% confidence intervals in Table 3 were calculated from 30 trial-specific importance estimates and do not capture every source of model-development uncertainty. Patient-level alignment was assessed using cosine similarity across unique trial pairs and a whole-trial patient-correspondence randomization procedure. This approach preserves trial-level attribution structure but remains an internal measure of alignment and requires confirmation on independently trained models and external patients.

The subgroup analysis adjusts one feature’s marginal distribution at a time and does not remove multivariable differences between patient groups. Its nominal weighted Student’s *t* tests compare each reweighted subgroup with a pooled evaluation reference that includes the subgroup itself. They also do not propagate uncertainty from estimating the feature-specific weights, and the one-sided tail was evaluated in the direction of the observed difference. These results are therefore exploratory. The absence of false-discovery-rate-adjusted findings and the small sizes of several subgroups further limit subgroup-specific interpretation.

Finally, model-specific SHAP explainers operated on the implemented prediction functions, and their outputs were not transformed to a common probability or log-odds scale. Normalization made relative attribution magnitudes comparable for aggregation, but the resulting performance-weighted profile is an empirical summary of model-specific attributions rather than the exact SHAP decomposition of a single probabilistic ensemble. SHAP describes fitted-model behavior and does not independently identify causal effects. Future work should use a fixed external evaluation cohort, preserve patient identifiers across trials, define a clinically meaningful prediction time, explain a common mortality-probability output across algorithms, and evaluate whether the aggregated attribution profile generalizes to other clinical populations.

## 6 Conclusions

This study integrates normalized performance-weighted ensemble SHAP aggregation with repeated training trials and distribution-adjusted subgroup analysis to interpret COVID-19 mortality predictions. Algorithms with similar predictive performance produced different attribution profiles, supporting the use of information from multiple fitted models when constructing a representative summary of feature importance.

The analysis distinguishes feature-importance magnitude, summarized across 30 trials using means and normal-approximation 95% confidence intervals, from patient-level attribution alignment, assessed using cosine similarity and patient-correspondence randomization. The aggregated profile identified clinically plausible patterns involving oxygenation, symptoms, laboratory measurements, and treatment context, while the subgroup comparisons generated exploratory patterns that did not remain below 0.05 after false-discovery-rate adjustment. These findings describe model-captured relationships rather than causal clinical effects and require validation using independent cohorts, a prespecified prediction time, and evaluation patients excluded from model training.

## Data Availability

Individual-level clinical data are not publicly available because they contain sensitive patient information and their use is restricted by institutional and ethical approvals. Aggregated results supporting the findings are provided in the manuscript and supplementary materials. Requests for access to de-identified data may be directed to the corresponding authors and will be considered subject to approval by the participating institutions and the relevant ethics oversight body.

## Supplementary Information

### Supplementary Figures

Supplementary Figures 1-16 display signed normalized ensemble SHAP attributions for the most important features within each clinical category. Each point represents one pooled evaluation observation after performance-weighted normalized attributions were averaged across the ten trials in its fixed evaluation batch. Positive values increase the normalized ensemble output relative to its baseline, whereas negative values decrease it. Color denotes the within-batch percentile rank of the observed feature value.

**Supplementary Figure 1:**
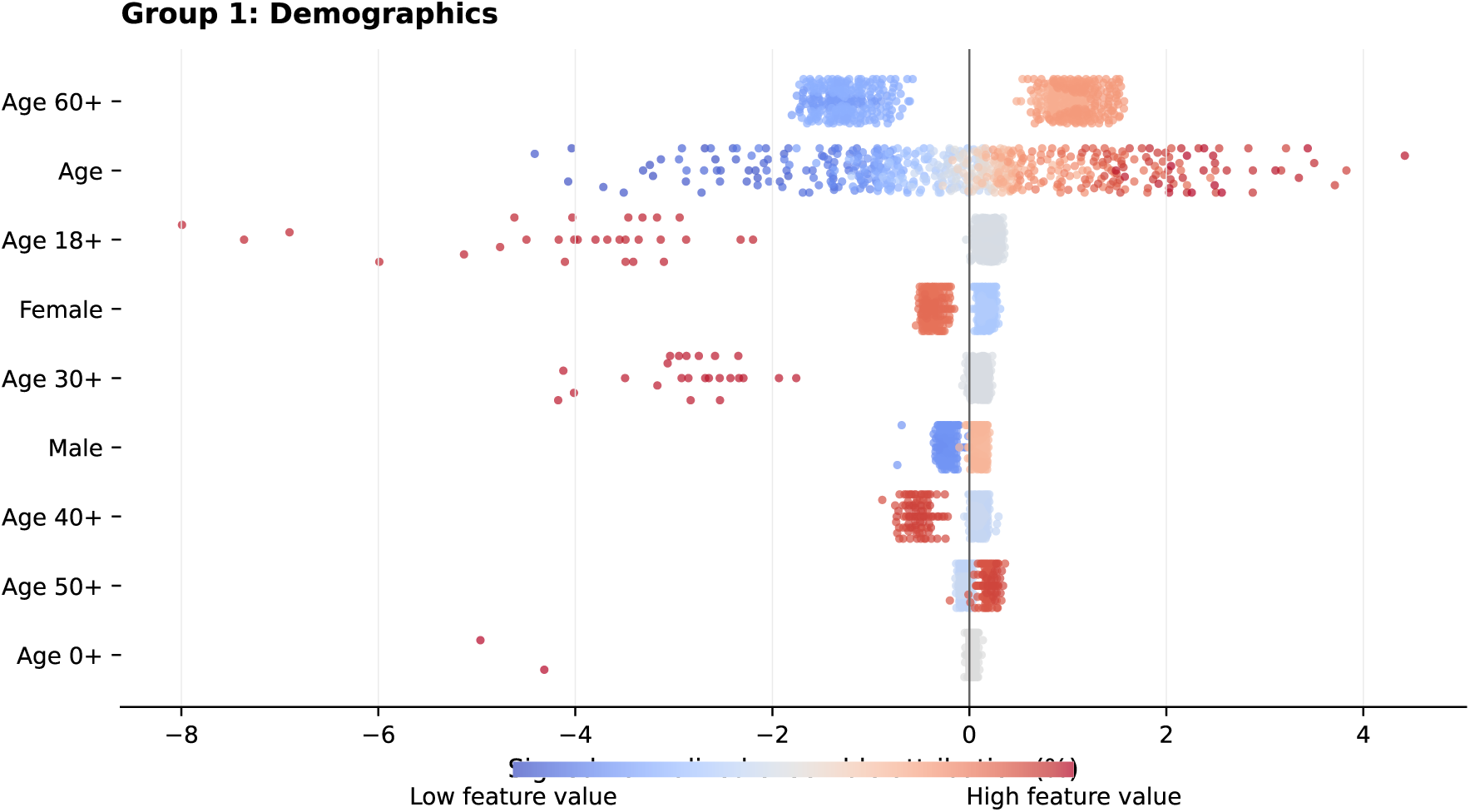
Signed normalized ensemble SHAP attributions for Group 1 (Demographics). The plot shows the 9 features with the greatest mean absolute importance in this category. Each point represents one evaluation observation after performance-weighted normalized attributions were averaged across the ten training trials in its fixed evaluation batch. Points to the right of zero increase the normalized ensemble output relative to its baseline; points to the left decrease it. Blue and red indicate lower and higher observed feature values, respectively. Numerical importance and alignment summaries are reported in main-manuscript Table 3.

**Supplementary Figure 2:**
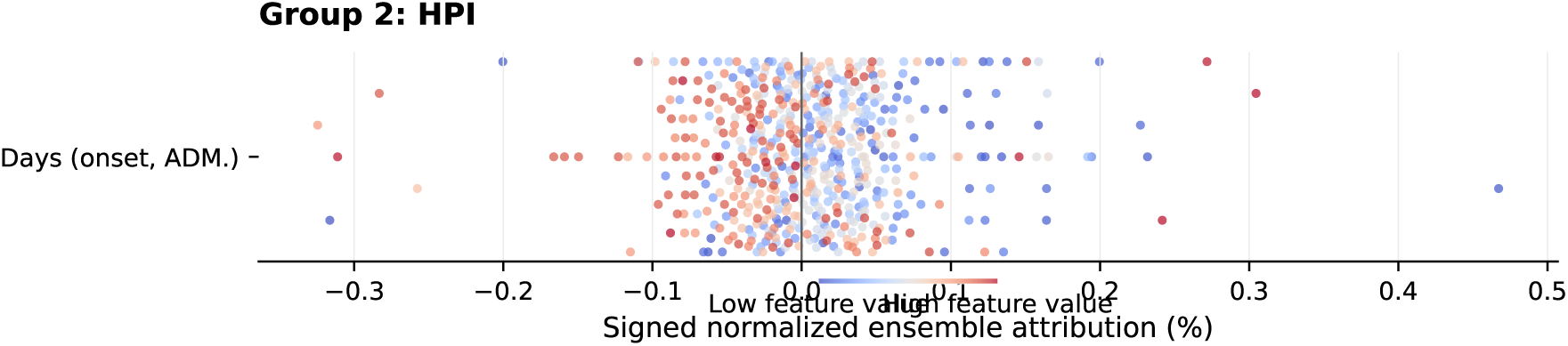
Signed normalized ensemble SHAP attributions for Group 2 (History of Present Illness (HPI)). The plot shows the 1 feature with the greatest mean absolute importance in this category. Each point represents one evaluation observation after performance-weighted normalized attributions were averaged across the ten training trials in its fixed evaluation batch. Points to the right of zero increase the normalized ensemble output relative to its baseline; points to the left decrease it. Blue and red indicate lower and higher observed feature values, respectively. Numerical importance and alignment summaries are reported in main-manuscript Table 3.

**Supplementary Figure 3:**
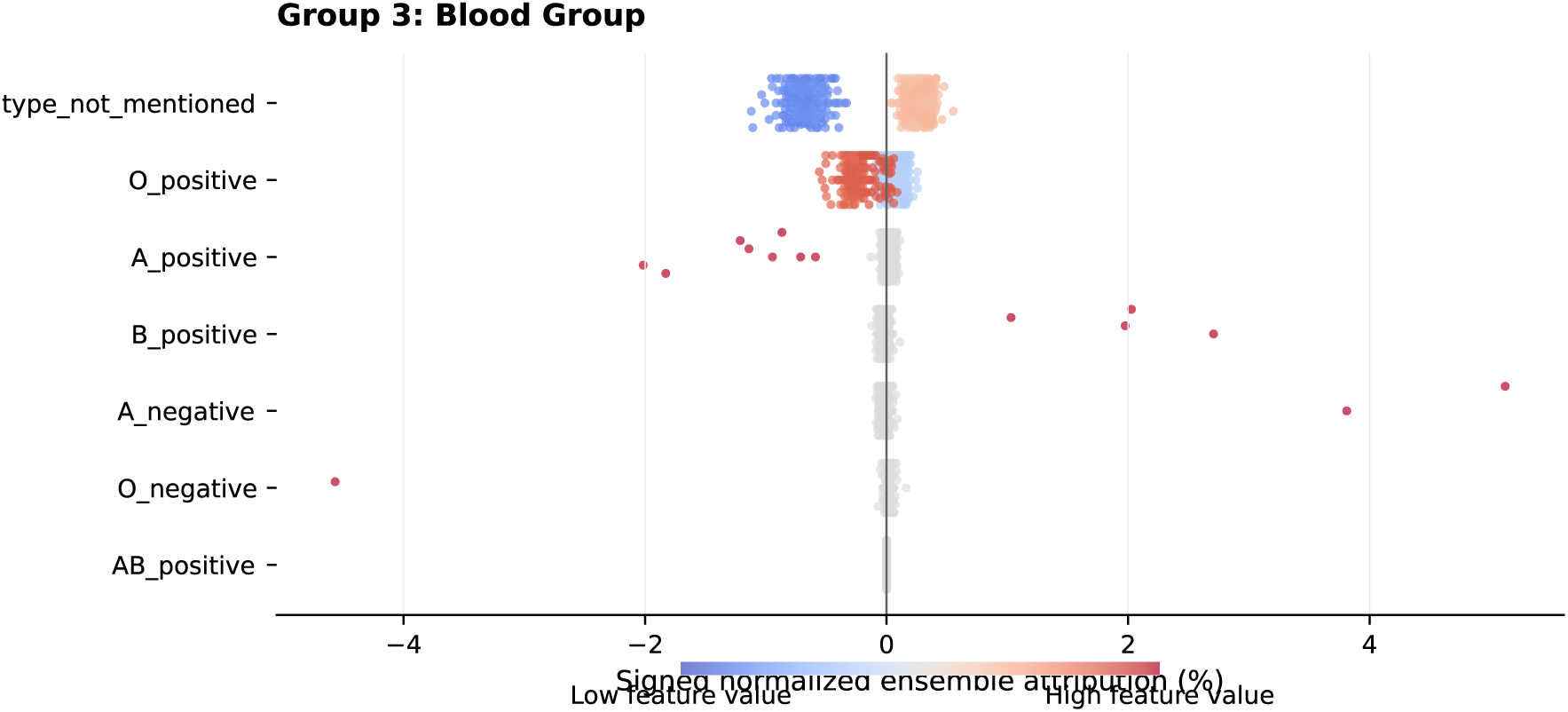
Signed normalized ensemble SHAP attributions for Group 3 (Blood Group). The plot shows the 7 features with the greatest mean absolute importance in this category. Each point represents one evaluation observation after performance-weighted normalized attributions were averaged across the ten training trials in its fixed evaluation batch. Points to the right of zero increase the normalized ensemble output relative to its baseline; points to the left decrease it. Blue and red indicate lower and higher observed feature values, respectively. Numerical importance and alignment summaries are reported in main-manuscript Table 3.

**Supplementary Figure 4:**
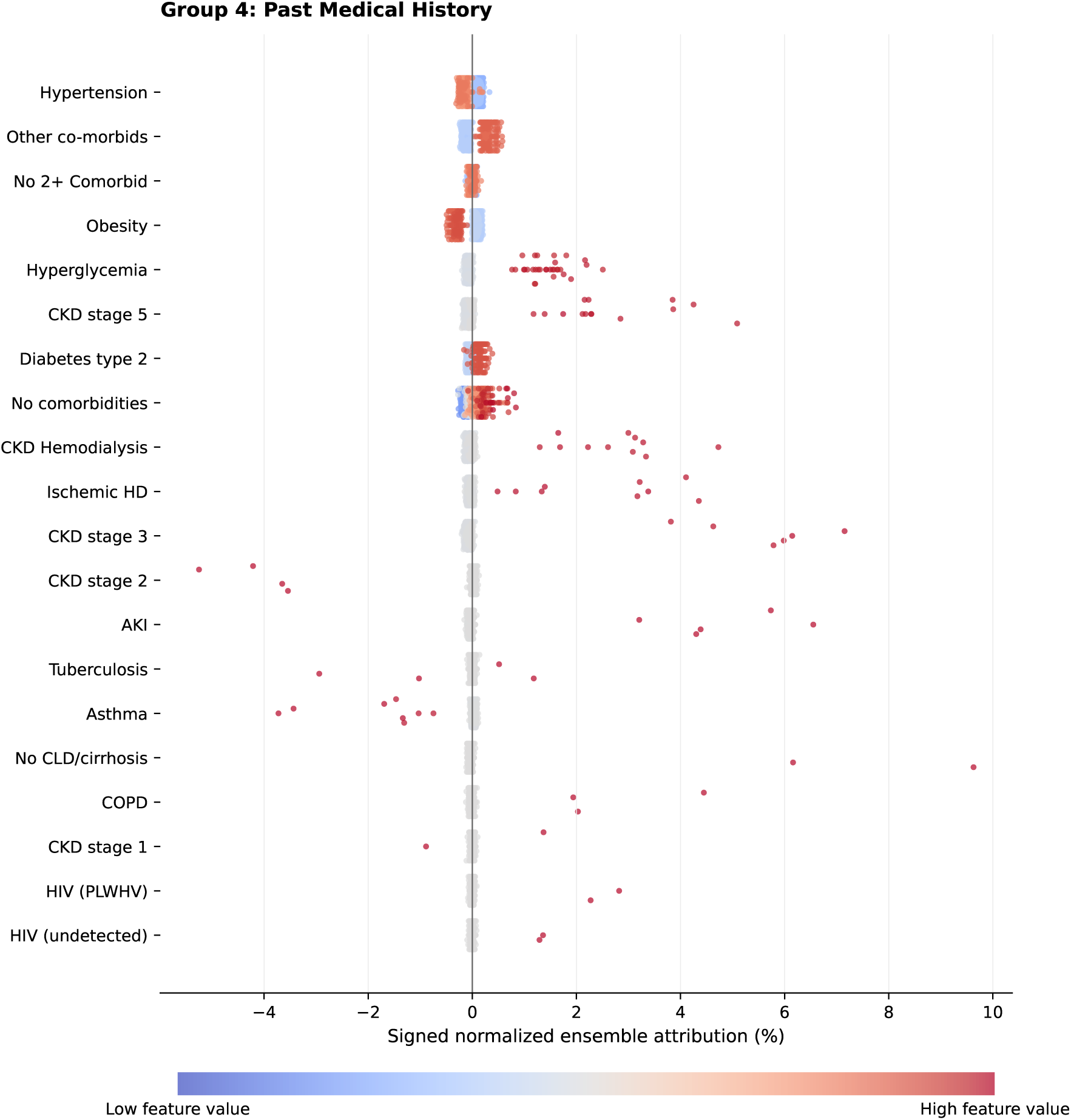
Signed normalized ensemble SHAP attributions for Group 4 (Past Medical History). The plot shows the 20 features with the greatest mean absolute importance in this category. Each point represents one evaluation observation after performance-weighted normalized attributions were averaged across the ten training trials in its fixed evaluation batch. Points to the right of zero increase the normalized ensemble output relative to its baseline; points to the left decrease it. Blue and red indicate lower and higher observed feature values, respectively. Numerical importance and alignment summaries are reported in main-manuscript Table 3.

**Supplementary Figure 5:**
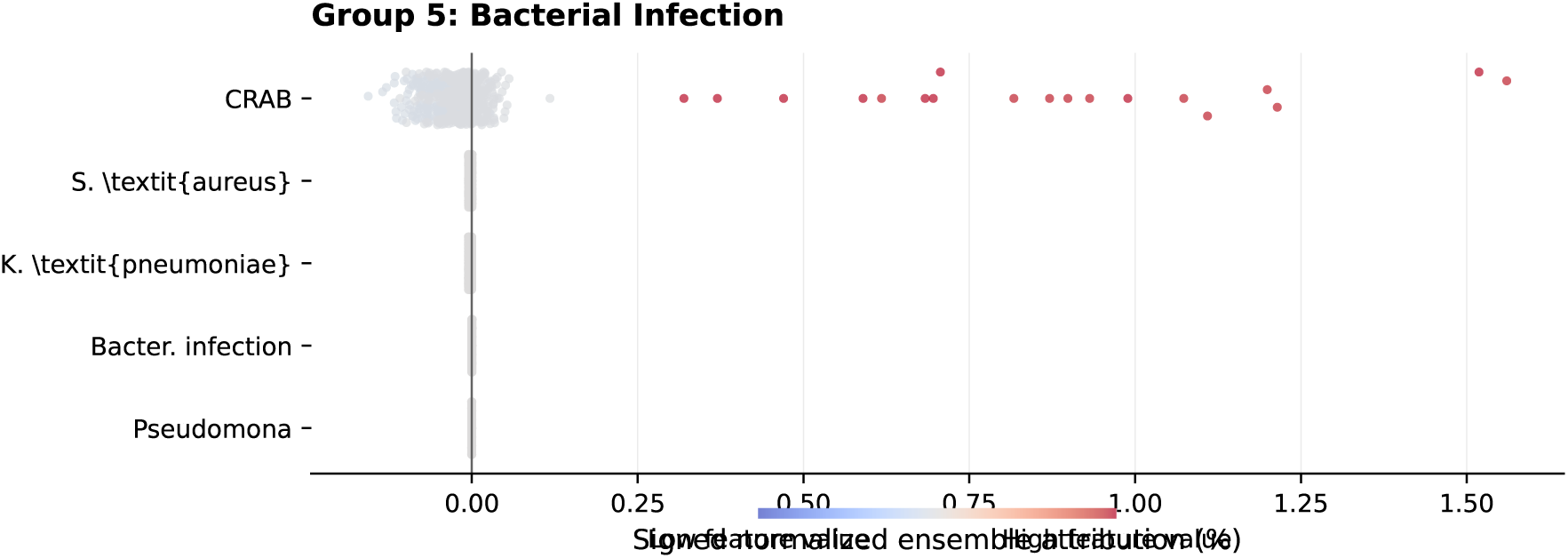
Signed normalized ensemble SHAP attributions for Group 5 (Bacterial Infection). The plot shows the 5 features with the greatest mean absolute importance in this category. Each point represents one evaluation observation after performance-weighted normalized attributions were averaged across the ten training trials in its fixed evaluation batch. Points to the right of zero increase the normalized ensemble output relative to its baseline; points to the left decrease it. Blue and red indicate lower and higher observed feature values, respectively. Numerical importance and alignment summaries are reported in main-manuscript Table 3.

**Supplementary Figure 6:**
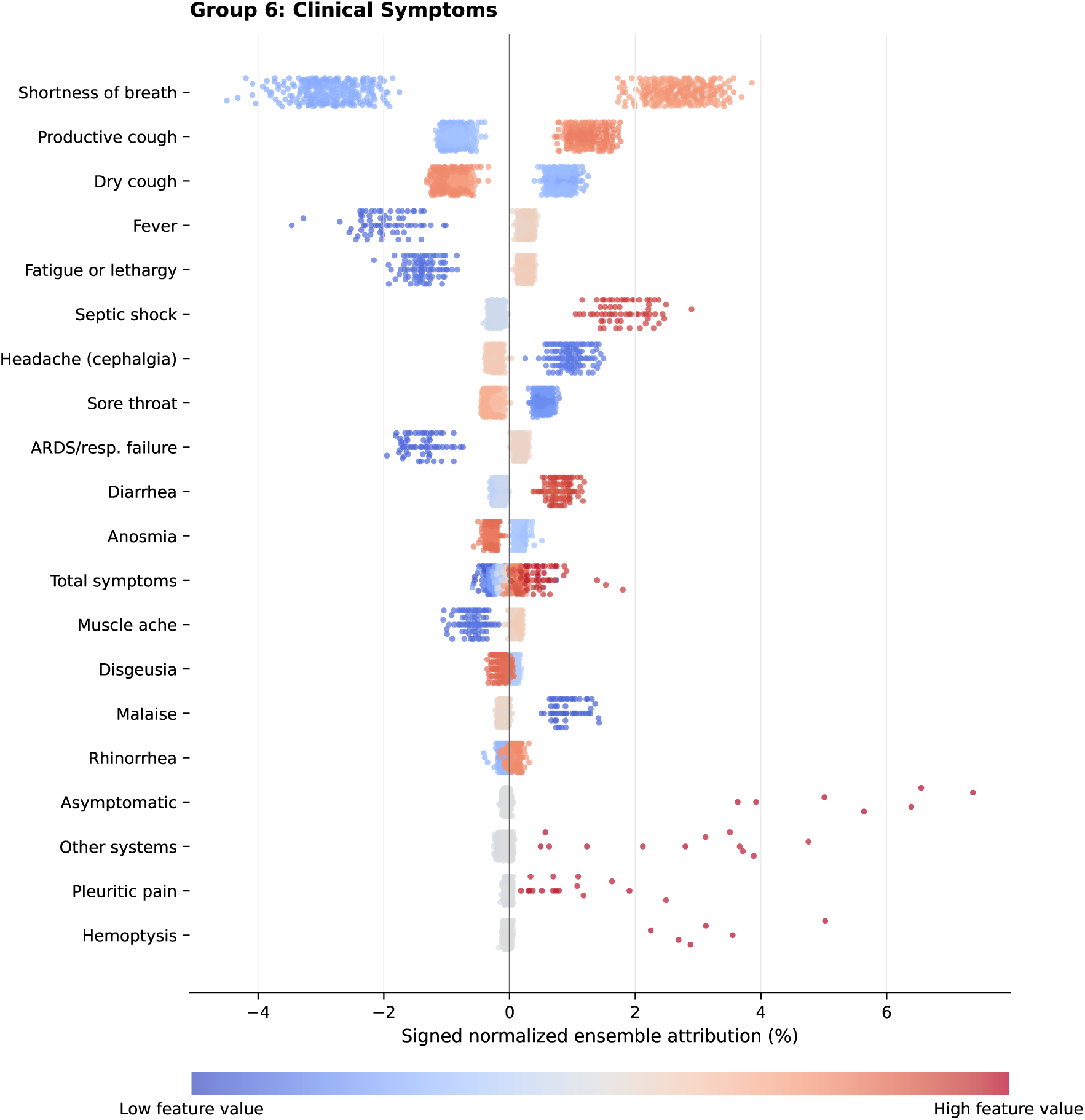
Signed normalized ensemble SHAP attributions for Group 6 (Clinical Symptoms). The plot shows the 20 features with the greatest mean absolute importance in this category. Each point represents one evaluation observation after performance-weighted normalized attributions were averaged across the ten training trials in its fixed evaluation batch. Points to the right of zero increase the normalized ensemble output relative to its baseline; points to the left decrease it. Blue and red indicate lower and higher observed feature values, respectively. Numerical importance and alignment summaries are reported in main-manuscript Table 3.

**Supplementary Figure 7:**
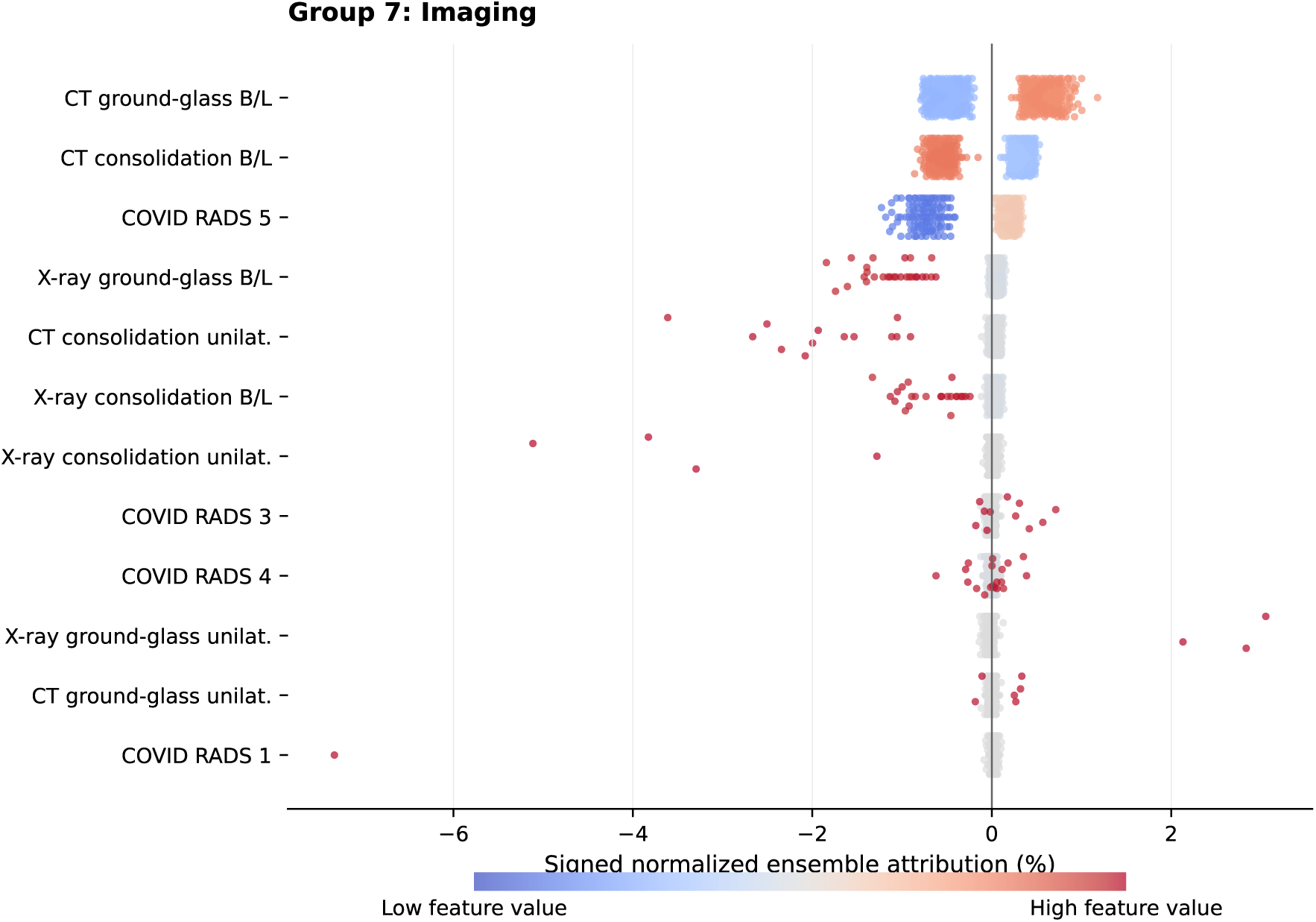
Signed normalized ensemble SHAP attributions for Group 7 (Imaging). The plot shows the 12 features with the greatest mean absolute importance in this category. Each point represents one evaluation observation after performance-weighted normalized attributions were averaged across the ten training trials in its fixed evaluation batch. Points to the right of zero increase the normalized ensemble output relative to its baseline; points to the left decrease it. Blue and red indicate lower and higher observed feature values, respectively. Numerical importance and alignment summaries are reported in main-manuscript Table 3.

**Supplementary Figure 8:**
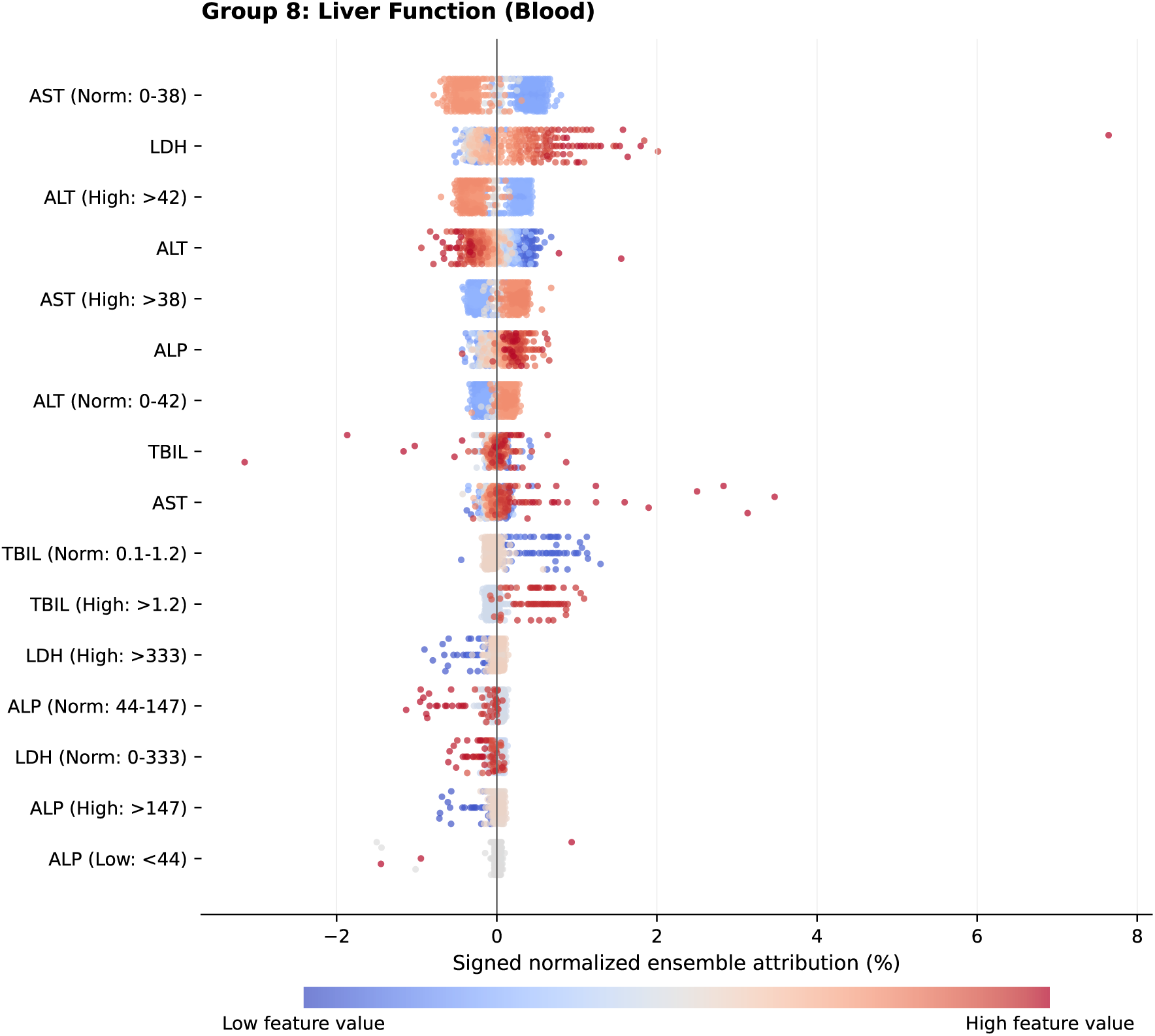
Signed normalized ensemble SHAP attributions for Group 8 (Liver Function Tests). The plot shows the 16 features with the greatest mean absolute importance in this category. Each point represents one evaluation observation after performance-weighted normalized attributions were averaged across the ten training trials in its fixed evaluation batch. Points to the right of zero increase the normalized ensemble output relative to its baseline; points to the left decrease it. Blue and red indicate lower and higher observed feature values, respectively. Numerical importance and alignment summaries are reported in main-manuscript Table 3.

**Supplementary Figure 9:**
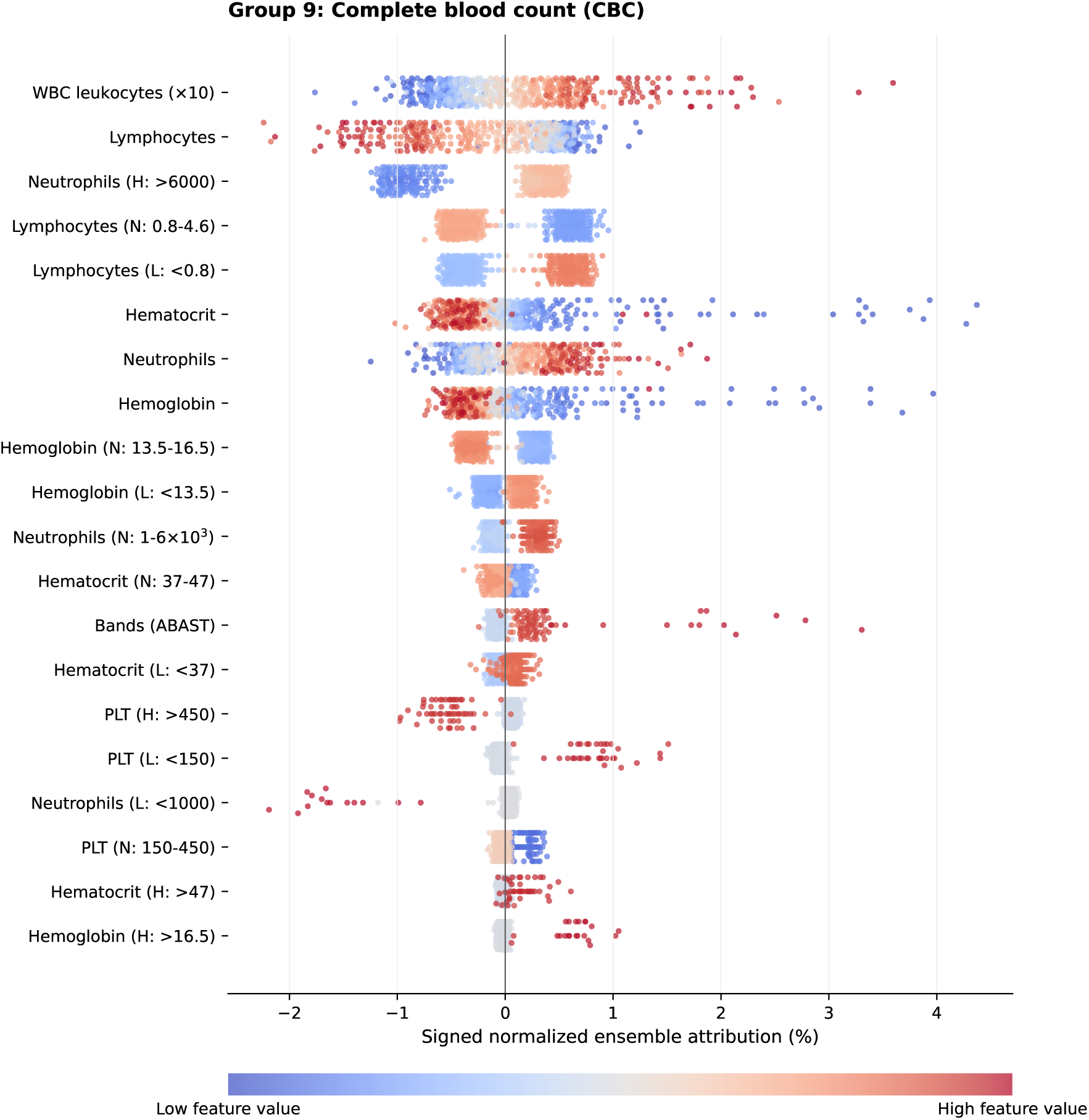
Signed normalized ensemble SHAP attributions for Group 9 (Complete Blood Count (CBC)). The plot shows the 20 features with the greatest mean absolute importance in this category. Each point represents one evaluation observation after performance-weighted normalized attributions were averaged across the ten training trials in its fixed evaluation batch. Points to the right of zero increase the normalized ensemble output relative to its baseline; points to the left decrease it. Blue and red indicate lower and higher observed feature values, respectively. Numerical importance and alignment summaries are reported in main-manuscript Table 3.

**Supplementary Figure 10:**
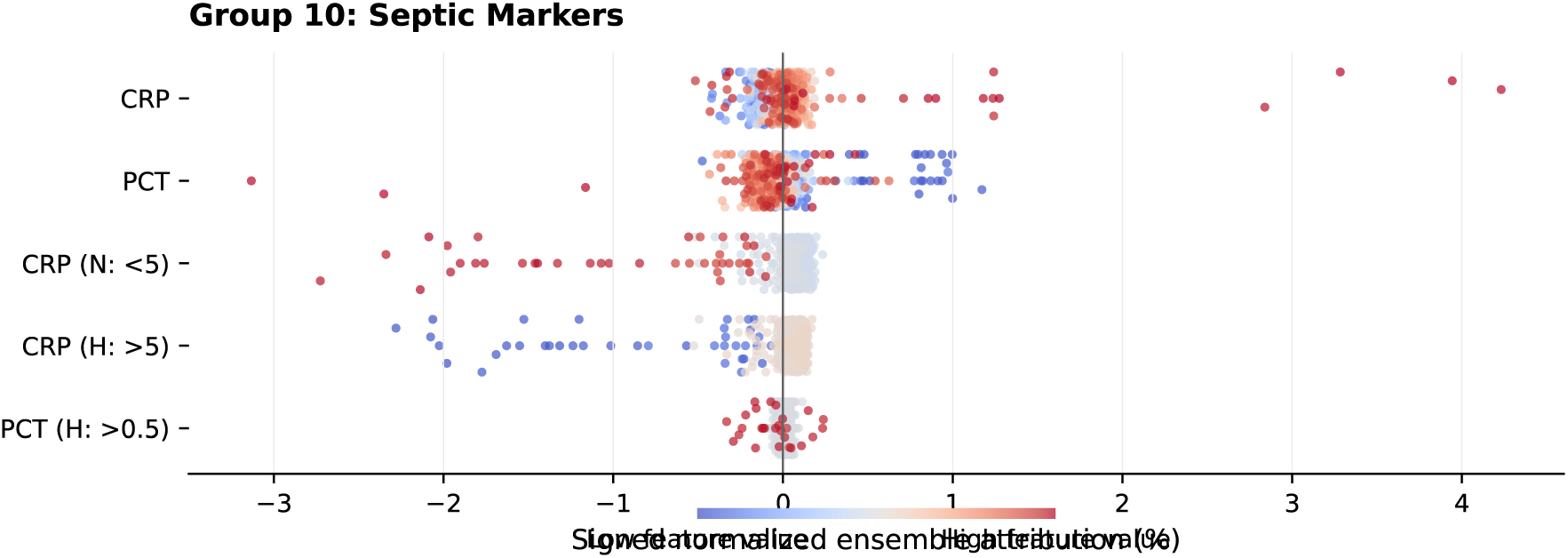
Signed normalized ensemble SHAP attributions for Group 10 (Septic Markers). The plot shows the 5 features with the greatest mean absolute importance in this category. Each point represents one evaluation observation after performance-weighted normalized attributions were averaged across the ten training trials in its fixed evaluation batch. Points to the right of zero increase the normalized ensemble output relative to its baseline; points to the left decrease it. Blue and red indicate lower and higher observed feature values, respectively. Numerical importance and alignment summaries are reported in main-manuscript Table 3.

**Supplementary Figure 11:**
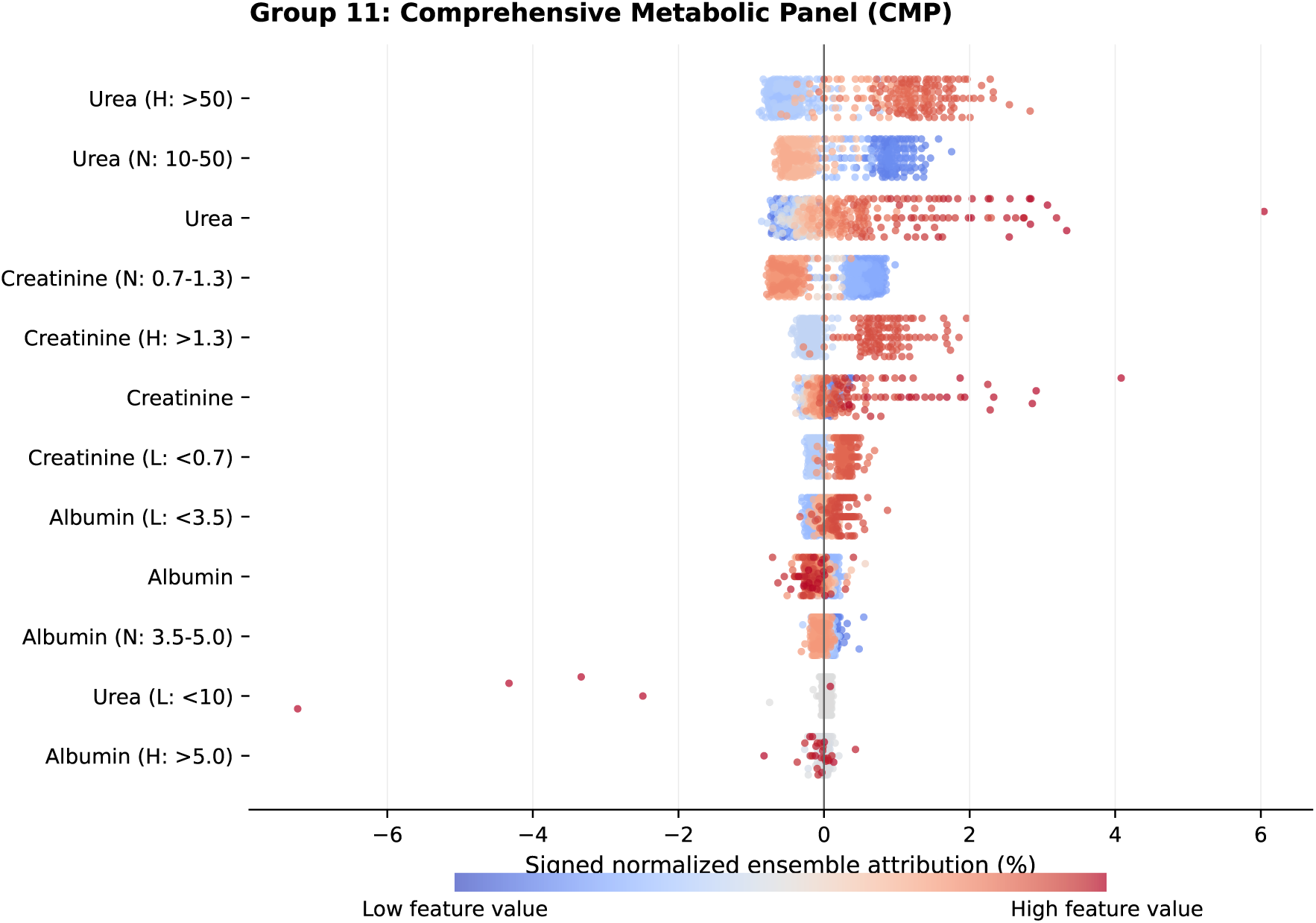
Signed normalized ensemble SHAP attributions for Group 11 (Comprehensive Metabolic Panel (CMP)). The plot shows the 12 features with the greatest mean absolute importance in this category. Each point represents one evaluation observation after performance-weighted normalized attributions were averaged across the ten training trials in its fixed evaluation batch. Points to the right of zero increase the normalized ensemble output relative to its baseline; points to the left decrease it. Blue and red indicate lower and higher observed feature values, respectively. Numerical importance and alignment summaries are reported in main-manuscript Table 3.

**Supplementary Figure 12:**
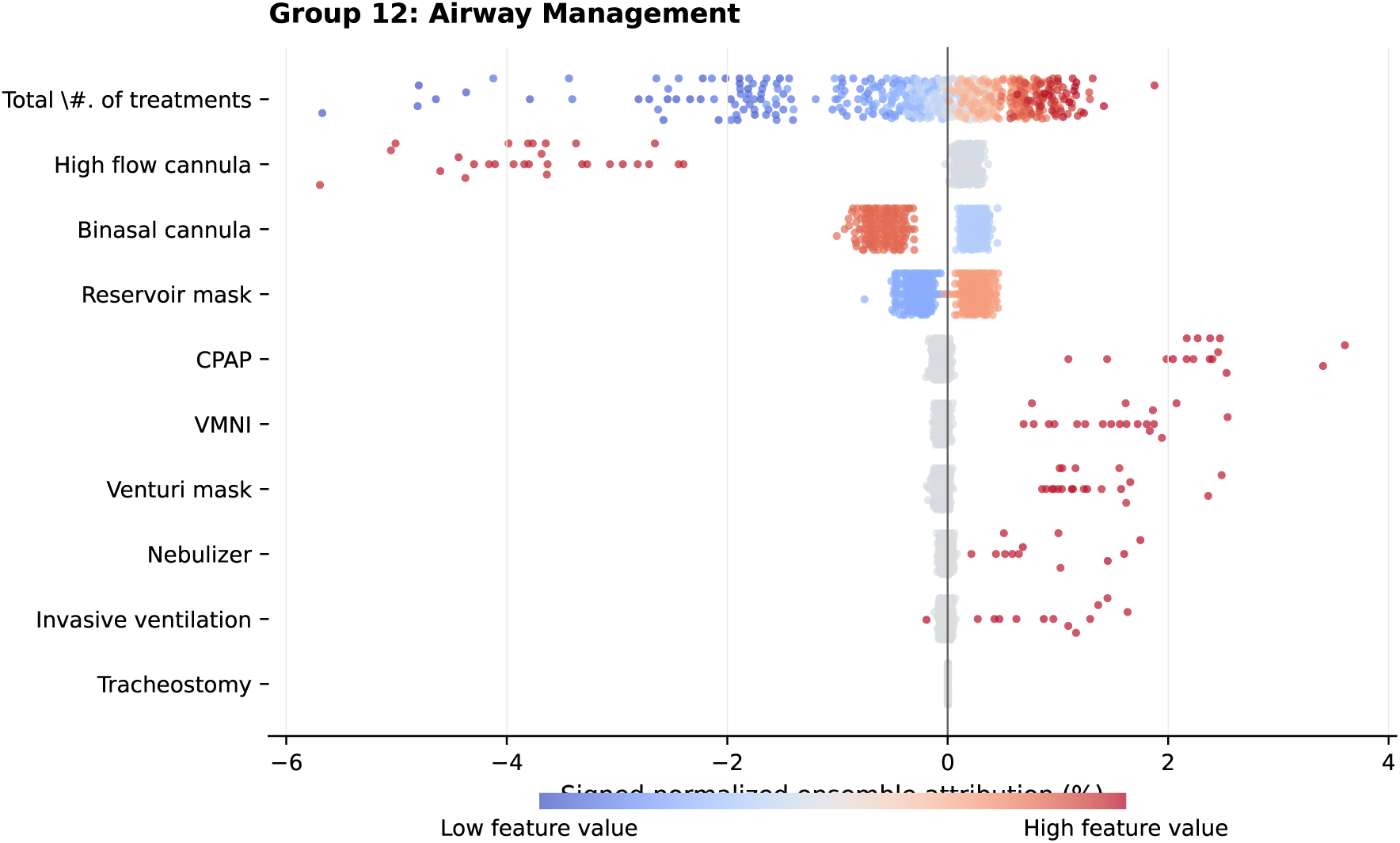
Signed normalized ensemble SHAP attributions for Group 12 (Airway Management). The plot shows the 10 features with the greatest mean absolute importance in this category. Each point represents one evaluation observation after performance-weighted normalized attributions were averaged across the ten training trials in its fixed evaluation batch. Points to the right of zero increase the normalized ensemble output relative to its baseline; points to the left decrease it. Blue and red indicate lower and higher observed feature values, respectively. Numerical importance and alignment summaries are reported in main-manuscript Table 3.

**Supplementary Figure 13:**
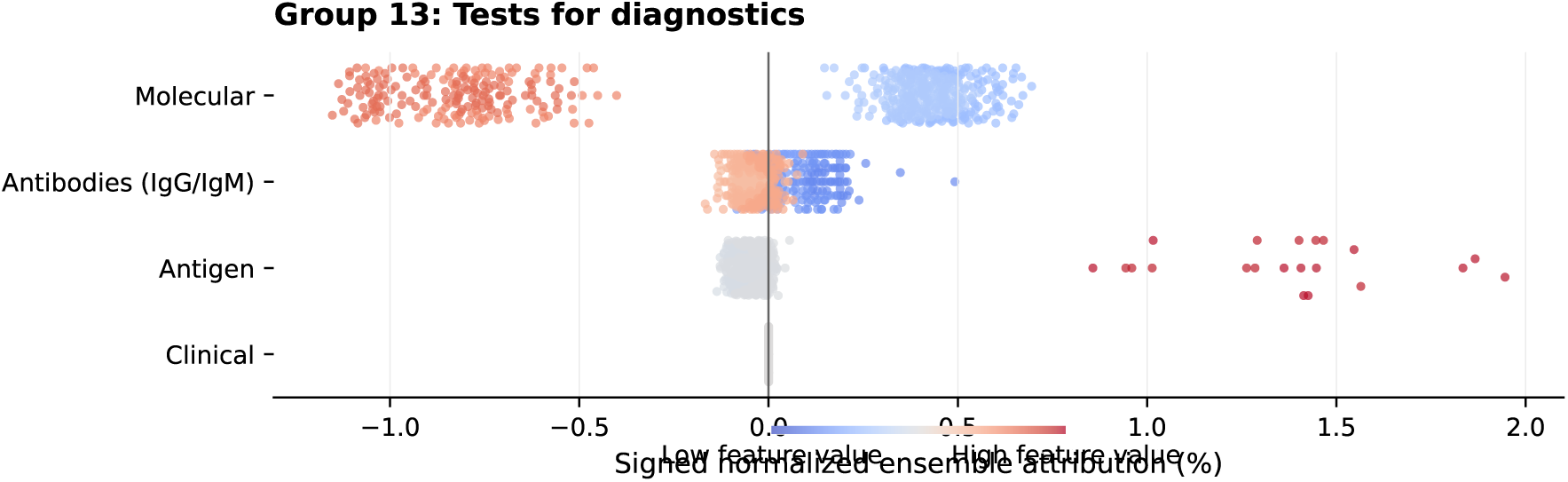
Signed normalized ensemble SHAP attributions for Group 13 (Diagnostic Tests). The plot shows the 4 features with the greatest mean absolute importance in this category. Each point represents one evaluation observation after performance-weighted normalized attributions were averaged across the ten training trials in its fixed evaluation batch. Points to the right of zero increase the normalized ensemble output relative to its baseline; points to the left decrease it. Blue and red indicate lower and higher observed feature values, respectively. Numerical importance and alignment summaries are reported in main-manuscript Table 3.

**Supplementary Figure 14:**
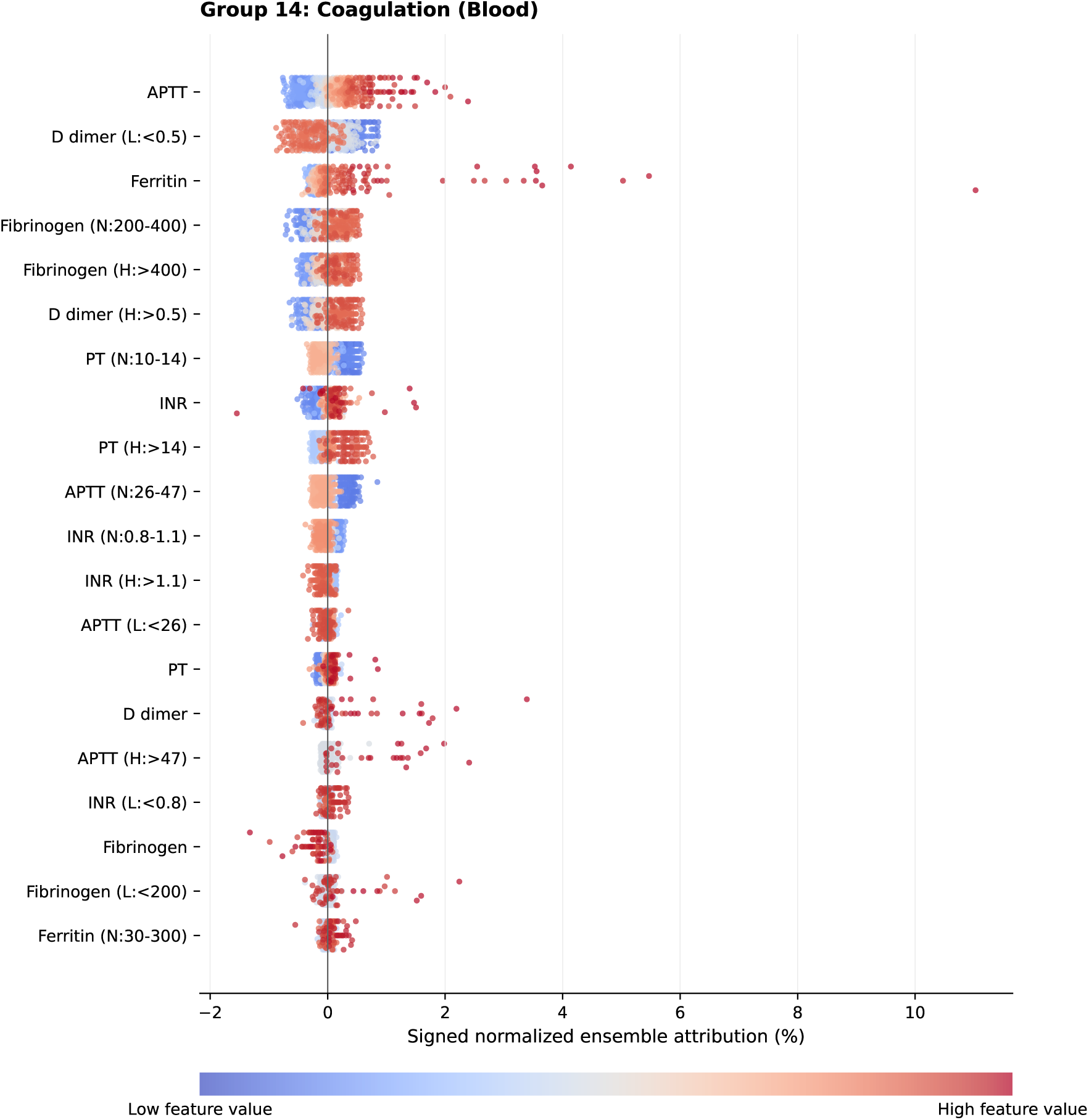
Signed normalized ensemble SHAP attributions for Group 14 (Coagulation). The plot shows the 20 features with the greatest mean absolute importance in this category. Each point represents one evaluation observation after performance-weighted normalized attributions were averaged across the ten training trials in its fixed evaluation batch. Points to the right of zero increase the normalized ensemble output relative to its baseline; points to the left decrease it. Blue and red indicate lower and higher observed feature values, respectively. Numerical importance and alignment summaries are reported in main-manuscript Table 3.

**Supplementary Figure 15:**
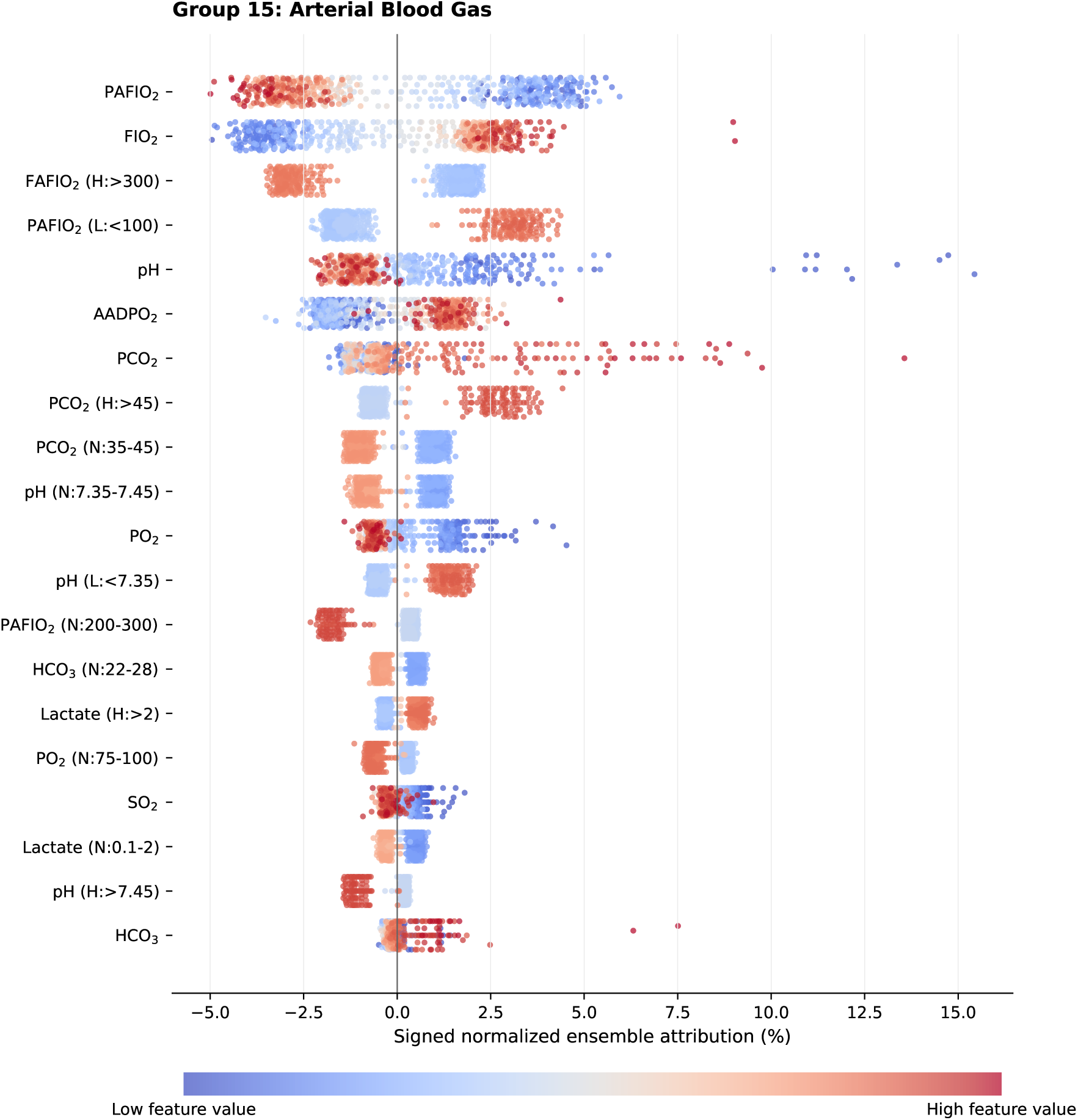
Signed normalized ensemble SHAP attributions for Group 15 (Arterial Blood Gas). The plot shows the 20 features with the greatest mean absolute importance in this category. Each point represents one evaluation observation after performance-weighted normalized attributions were averaged across the ten training trials in its fixed evaluation batch. Points to the right of zero increase the normalized ensemble output relative to its baseline; points to the left decrease it. Blue and red indicate lower and higher observed feature values, respectively. Numerical importance and alignment summaries are reported in main-manuscript Table 3.

**Supplementary Figure 16:**
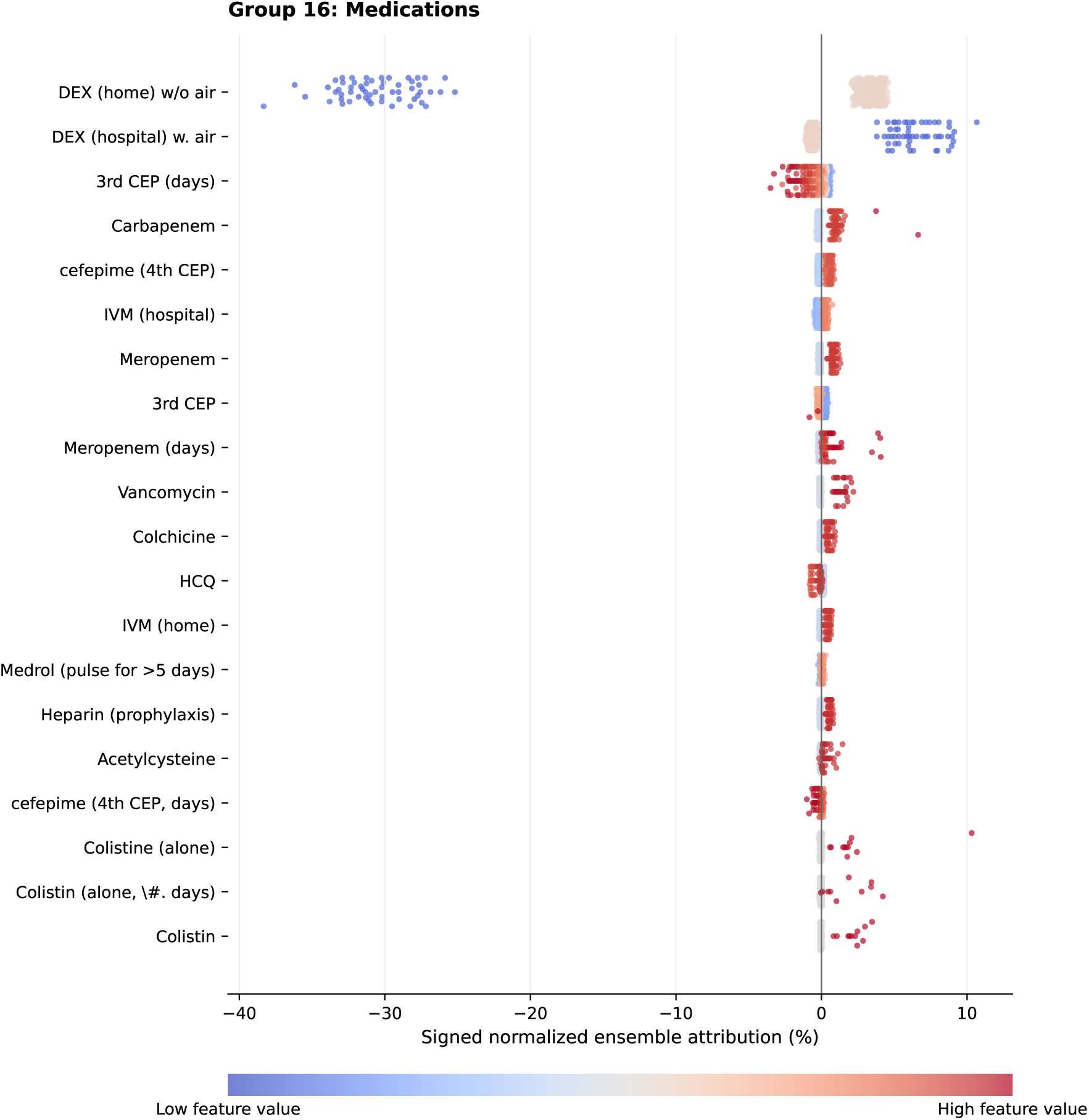
Signed normalized ensemble SHAP attributions for Group 16 (Medications). The plot shows the 20 features with the greatest mean absolute importance in this category. Each point represents one evaluation observation after performance-weighted normalized attributions were averaged across the ten training trials in its fixed evaluation batch. Points to the right of zero increase the normalized ensemble output relative to its baseline; points to the left decrease it. Blue and red indicate lower and higher observed feature values, respectively. Numerical importance and alignment summaries are reported in main-manuscript Table 3.

### Supplementary Tables

#### Condensed Subgroup Results

The complete results for all 3,113 subgroup–feature comparisons, including both Benjamini–Hochberg adjustments, are provided as Supplementary Data S1 in machine-readable CSV format. Tables 1 and 2 summarize inferential eligibility and the 64 comparisons with nominal one-sided *p <* 0.05, respectively. No comparison remained below 0.05 after adjustment within subgroup or across all eligible comparisons.

**Supplementary Table 1:** Summary of inferential eligibility and multiplicity adjustment by subgroup.

| Subgroup | $n_G$ | Total | Eligible | Small $n$ | Zero var. | KS > 0.10 | Defining | Nominal $p < 0.05$ | Min. $p$ | Min. $q_{\text{within}}$ | Min. $q_{\text{global}}$ |
| --- | --- | --- | --- | --- | --- | --- | --- | --- | --- | --- | --- |
| Age $\geq 50$ years | 109 | 283 | 257 | 0 | 19 | 4 | 3 | 8 | 0.017 | 0.499 | 0.499 |
| Male | 374 | 283 | 262 | 0 | 19 | 2 | 0 | 0 | 0.059 | 0.499 | 0.499 |
| Female | 177 | 283 | 262 | 0 | 19 | 2 | 0 | 12 | 0.010 | 0.498 | 0.499 |
| Blood type A | 10 | 283 | 0 | 283 | 0 | 0 | 0 | 0 | – | – | – |
| Blood type B | 4 | 283 | 0 | 283 | 0 | 0 | 0 | 0 | – | – | – |
| Blood type O | 143 | 283 | 257 | 0 | 19 | 2 | 5 | 7 | 0.004 | 0.496 | 0.499 |
| Asthma or COPD | 11 | 283 | 0 | 283 | 0 | 0 | 0 | 0 | – | – | – |
| Metabolic or cardiovascular disease | 303 | 283 | 258 | 0 | 19 | 5 | 1 | 4 | 0.023 | 0.497 | 0.499 |
| Kidney disease | 30 | 283 | 209 | 0 | 19 | 53 | 2 | 18 | < 0.001 | 0.153 | 0.499 |
| Chronic viral or liver disease | 4 | 283 | 0 | 283 | 0 | 0 | 0 | 0 | – | – | – |
| At least two documented comorbidities | 214 | 283 | 258 | 0 | 19 | 6 | 0 | 15 | 0.002 | 0.467 | 0.499 |
| Total | – | 3,113 | 1,763 | 1,132 | 133 | 74 | 11 | 64 | < 0.001 | 0.153 | 0.499 |

Notes. Total is the number of encoded features evaluated descriptively in each subgroup. Eligible denotes comparisons included in inferential testing. Small $n$ denotes subgroups containing fewer than 20 evaluation observations; Zero var. denotes zero patient-level attribution variance; KS > 0.10 denotes inadequate post-reweighting balance; Defining denotes features used to define the subgroup. Min. $q_{\text{within}}$ is the smallest Benjamini-Hochberg-adjusted value within the subgroup, and Min. $q_{\text{global}}$ is the smallest adjusted value under correction across all 1,763 eligible comparisons.

**Supplementary Table 2:**
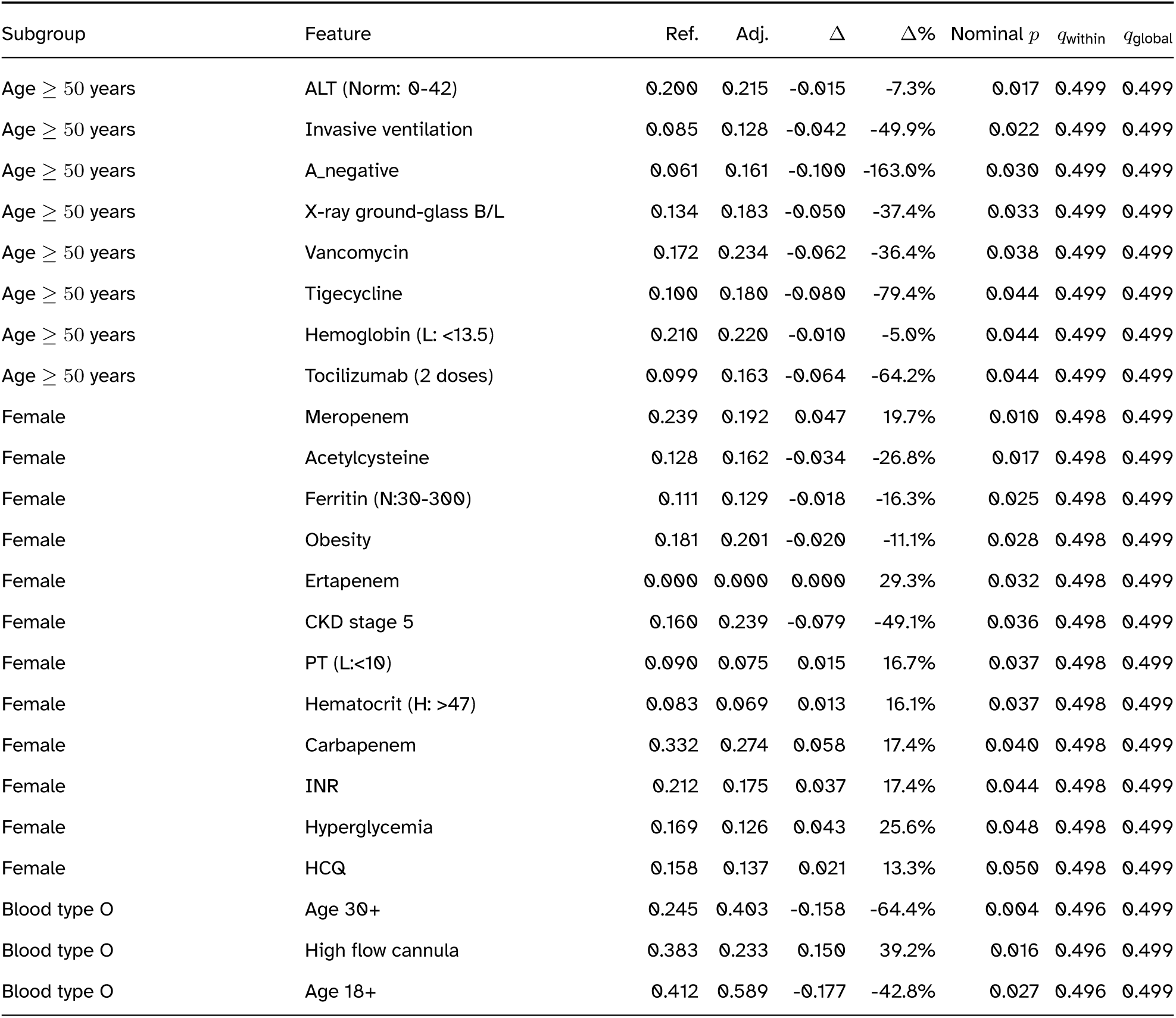
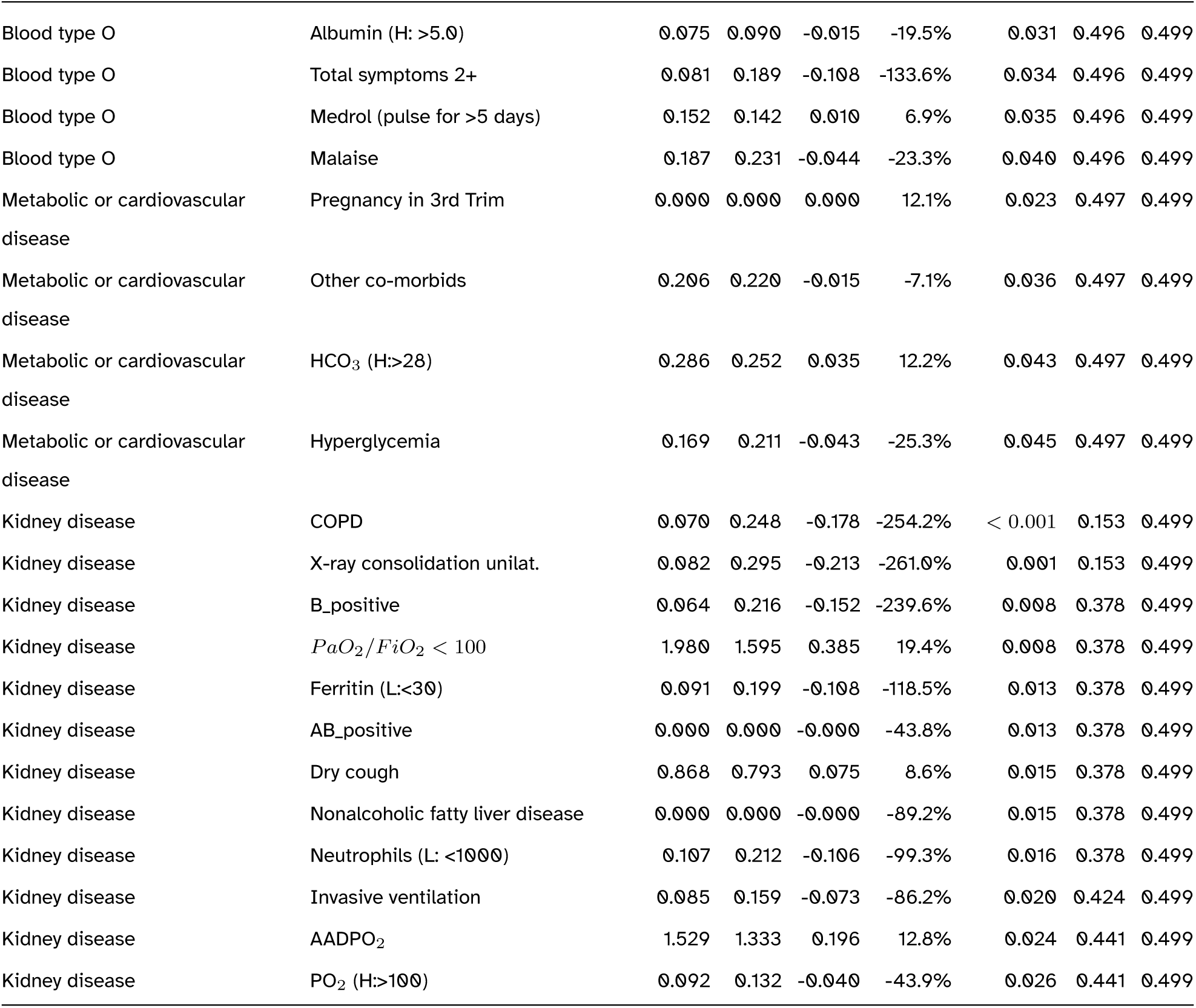
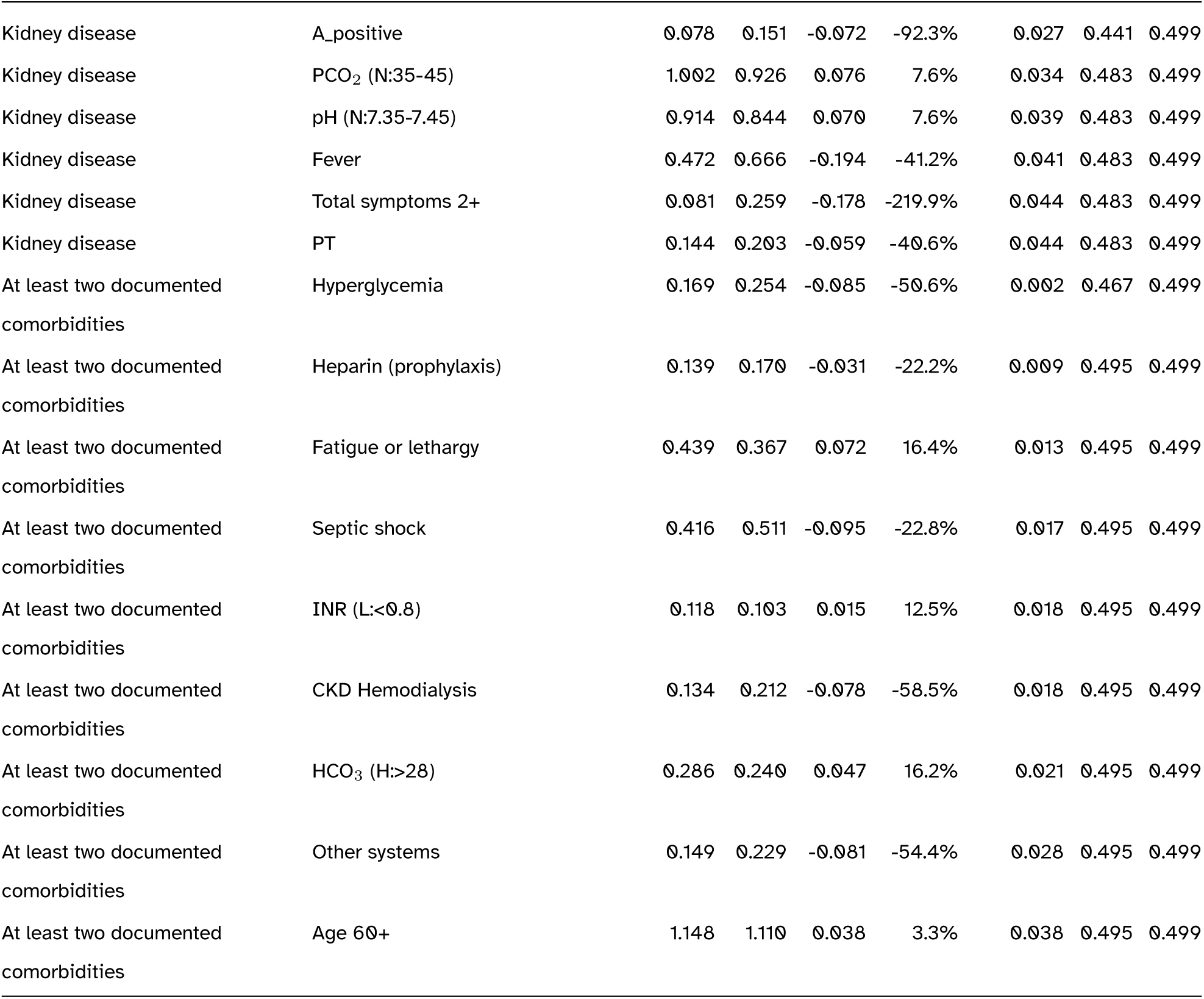
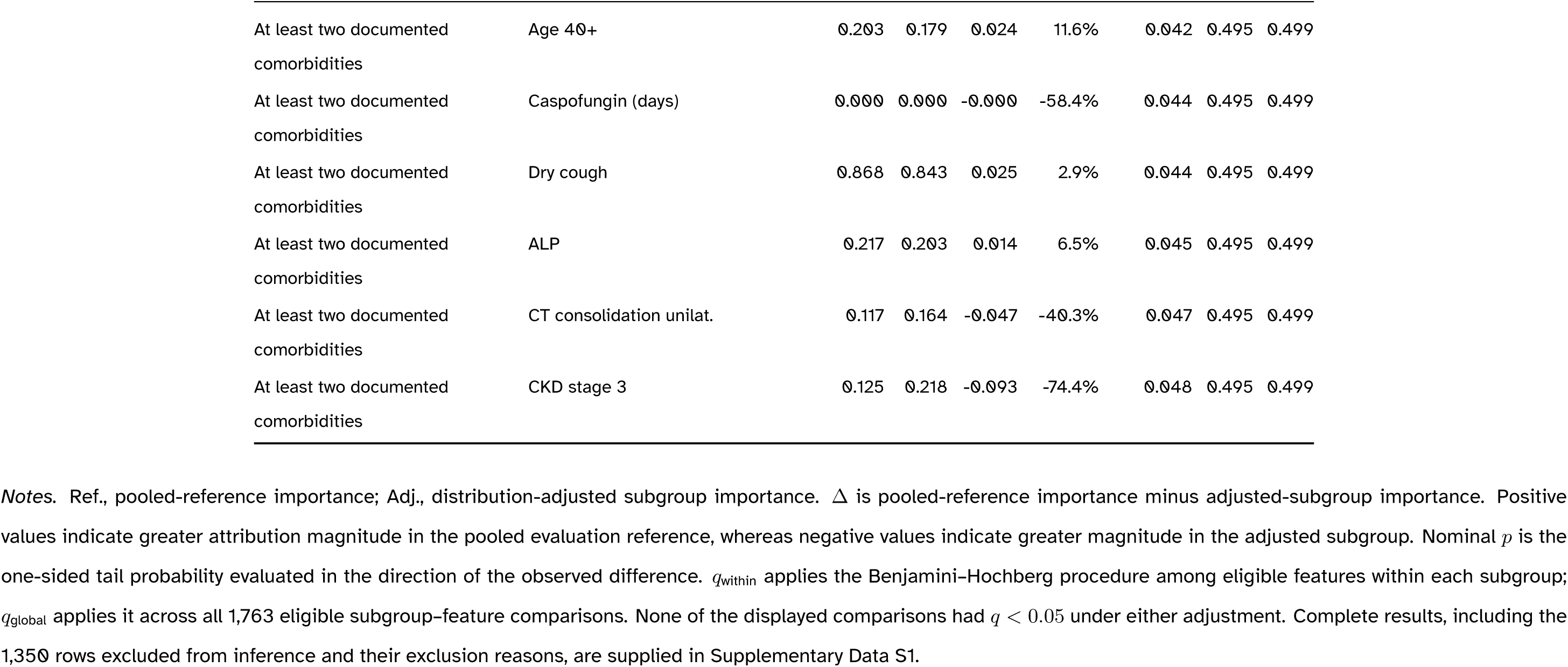
Subgroup-feature comparisons with nominal one-sided *p <* 0.05.

## Notes

### Competing Interest Statement

The authors have declared no competing interest.

### Author Declarations

The Comite de Investigacion y Etica, Oficina de Capacitacion, Investigacion y Docencia, Red Asistencial La Libertad, EsSalud, Peru, approved the use and analysis of the clinical data for this study in 2023 (approval number: 87CIYE-O.C.I.YD-RALL-ESSALUD-2023). Written informed consent for the collection and research use of clinical data was obtained from all participants.

